# The impact of retracted randomised controlled trials on systematic reviews and clinical practice guidelines: a meta-epidemiological study

**DOI:** 10.1101/2022.01.30.22270124

**Authors:** Yuki Kataoka, Masahiro Banno, Yasushi Tsujimoto, Takashi Ariie, Shunsuke Taito, Tomoharu Suzuki, Shiho Oide, Toshi A. Furukawa

## Abstract

**Objectives:** To investigate whether and when the correction is done in Systematic Reviews (SRs) and Clinical Practice Guidelines (CPGs) when their included Randomised Controlled Trials (RCTs) have been retracted.

**Design:** A meta-epidemiological study.

**Data sources:** The Retraction Watch Database.

**Eligibility criteria for selecting studies:** SRs and CPGs citing the retracted RCTs on Web of Science.

**Review methods:** We investigated how often the retracted RCTs were cited in SRs and CPGs. We also investigated whether and when such SRs and CPGs corrected themselves by visually inspecting their current web pages. We summarized the proportion of correction and the time from retraction to correction.

**Results:** We identified 98 retracted RCTs as well as 360 articles (335 SRs and 25 CPGs) citing them. Among the 360 articles, 157 (44%) were published after the retraction, 203 (56%) were published before retraction. Among 77 articles published citing already retracted RCTs in their evidence synthesis without caution, none corrected themselves after publication. Of 203 articles published before retraction, 149 included RCTs that were later retracted in their evidence synthesis. Among them, one SR was retracted due to plagiarism. Only 5% of SRs (6/130) and 11% of CPGs (2/18) corrected their results.

**Conclusions:** A large number of SRs and CPGs included already retracted RCTs without caution and never corrected themselves. When SRs and CPGs had included RCTs which were later retracted, only a small minority corrected their evidence syntheses. The scientific community, including publishers and researchers, should make systematic and concerted efforts to remove the impact of retracted RCTs.

**What is already known on this topic:**

- Systematic Reviews (SRs) and Clinical Practice Guidelines (CPGs) aggregating randomised controlled trials (RCTs) are important sources of information for clinical decision making.
- There are anecdotal reports of publications citing retracted RCTs and point to the problem of their continued citation after retraction.
- However, there are no studies that comprehensively examined the fate of retracted RCTs on SRs and CPGs in their evidence synthesis.

**What this study adds:**

- A considerable number of SRs and CPGs cited already retracted RCTs and none corrected themselves later.
- Only a small minority of SRs (5%, 6/130) and CPGs (11%, 2/18) which cited RCTs that were later retracted corrected their findings after the retraction was announced.
- The results indicate that publishers and researchers should make efforts to remove the impact of retracted RCT.

## Introduction

Systematic Reviews (SR) and Clinical Practice Guidelines (CPG) aggregating randomised controlled trials (RCT) are vital sources of information for clinical decision making [1]. There are guidelines on how to report SRs [2] and to create CPGs [3] in a rigorous scientific manner. One important point to remember is that all these methodologies assume that the data used in their evidence synthesis is valid [4].

In recent years, much attention has been paid to the retraction of papers due to scientific misconduct [5,6]. Notable recent examples include a RCT of ivermectin [7] and a cohort study of hydroxychloroquine, both for COVID-19 [8]. An increasing number of studies have investigated the fate of such retracted studies: some studies have evaluated the impact of retracted articles on social media [9] or reported on the ongoing citations of retracted articles in some specialties such as radiation oncology, dentistry or COVID-19 [6,10–12]. The Cochrane has developed a new policy to address potentially problematic studies including retraction [4]. The policy set out how we should manage the retracted articles.

However, to the best of the current authors’ knowledge, no studies have comprehensively examined how retracted RCTs managed in the evidence synthesis in SRs and CPGs. In this study we therefore investigated whether and when the correction is done in SRs and CPGs when they included retracted RCTs.

## Methods

### Protocol and registration

This meta-epidemiological study was conducted and reported in accordance with a guideline for reporting meta-epidemiological methodology research [13]. (Table S1) The protocol of the present study has been published in OSF [14].

### Search and selection for retracted RCTs

We searched the Retraction Watch Database (RWD) for retracted RCTs on 27^th^ July 2021 using the term “random*” [15]. RWD systematically retrieves retracted biomedical articles since 1756 including more than 28000 entries [15]. The database contains titles of articles but not abstracts. Two independent reviewers then identified RCTs from the full text articles retrieved based on the search results. An RCT is defined as “a work that reports on a clinical trial that involves at least one test treatment and one control treatment, concurrent enrollment and follow-up of the test- and control-treated groups, and in which the treatments to be administered are selected by a random process” [16]. We did not include quasi-randomised controlled trials. Disagreements were resolved through discussion.

### Search and identification of SRs and CPGs citing retracted RCTs

We searched each retracted RCT in the Web of Science (WOS) on 18^th^ Oct 2021 [17] to identify references which cited the retracted RCTs. The articles not indexed in the WOS were excluded. We placed no restrictions on date or language.

We included SRs and CPGs citing the identified retracted RCT articles. The SR was defined as “a scientific investigation that focuses on a specific question and uses explicit, prespecified scientific methods to identify, select, assess, and summarize the findings of similar but separate studies” [3]. We included any SR, with or without meta-analysis. The definition of the CPG was “statements that include recommendations intended to optimize patient care. They are informed by systematic reviews of evidence and an assessment of the benefits and harms of alternative care options” [3].

From the searched titles and abstracts, two independent reviewers selected SRs and CPGs. Disagreements were resolved through discussion. We then retrieved the full text articles and two independent reviewers finally identified SRs and CPGs. If necessary, a third reviewer acted as arbiter. We excluded articles published in journals without websites because judgement of correction of SR or CPG in such journals was difficult.

### Primary outcome

We calculated the proportion of correction among SRs and CPGs, respectively. For SRs, the numerator was the number of SR articles which corrected the results. The denominator was the number of SR articles which cited the retracted article in the results section for evidence synthesis. For CPGs, the numerator was the number of CPGs which corrected the explanation of recommendation considering the cited retracted RCT. We presented the outcome separately for articles published before and after retraction.

### Data extraction

We extracted the following data from the search results of the WOS of the included articles: the number of authors, country of the authors, publication date, journal impact factor in 2020, number of citations as of 13 ^th^ Nov 2021, funding (for-profit, non-profit, none, unclear), and research areas [18]. We used the selenium package version 3.141.0 and the ChromeDriver version 96.0 .4664.45 under python 3.7 to extract these data.

Two independent reviewers evaluated where the retracted articles were cited in the full text of SRs or CPGs. In addition, one reviewer inspected the web page of the included SR and CPG to determine whether their texts were corrected and if corrected, when they were corrected. Then another reviewer confirmed the results of the inspection. We resolved the disagreements through discussion.

### Statistical analysis

We used descriptive statistics to summarise. For survival analysis, we used the publication and retraction date of RCTs from RWD. We estimated time to correction using the Kaplan-Meier method. We used R ver. 4.1.2 (R Foundation for Statistical Computing, Vienna, Austria).

### Ethical consideration

We used publicly available data only, and ethical considerations for participants were unnecessary.

### Patient and public involvement

Patients and members of the public were not involved in the research because it was designed to investigate the current methodological practice in the SR and CPG.

### Differences between the protocol and the review

We did not conduct univariate and multivariable analyses, because of the small number of corrected SRs or CPGs.

## Results

### Search results

Figure 1 shows the study flow chart. From 28960 records indexed in the RWD, we idetified potentially eligible 114 RCT records. Excluding one protocol and 15 records not indexed in the WOS, we finally included 98 retracted RCT articles (Table S2). By citation search of the 98 retracted RCT articles in the WOS, we found 4822 records citing these retracted RCTs. By title and abstract screening, we selected 448 articles for full text examination. After full text screening, we finally included 335 SRs and 25 CPGs (Table S3).

**Figure 1.**
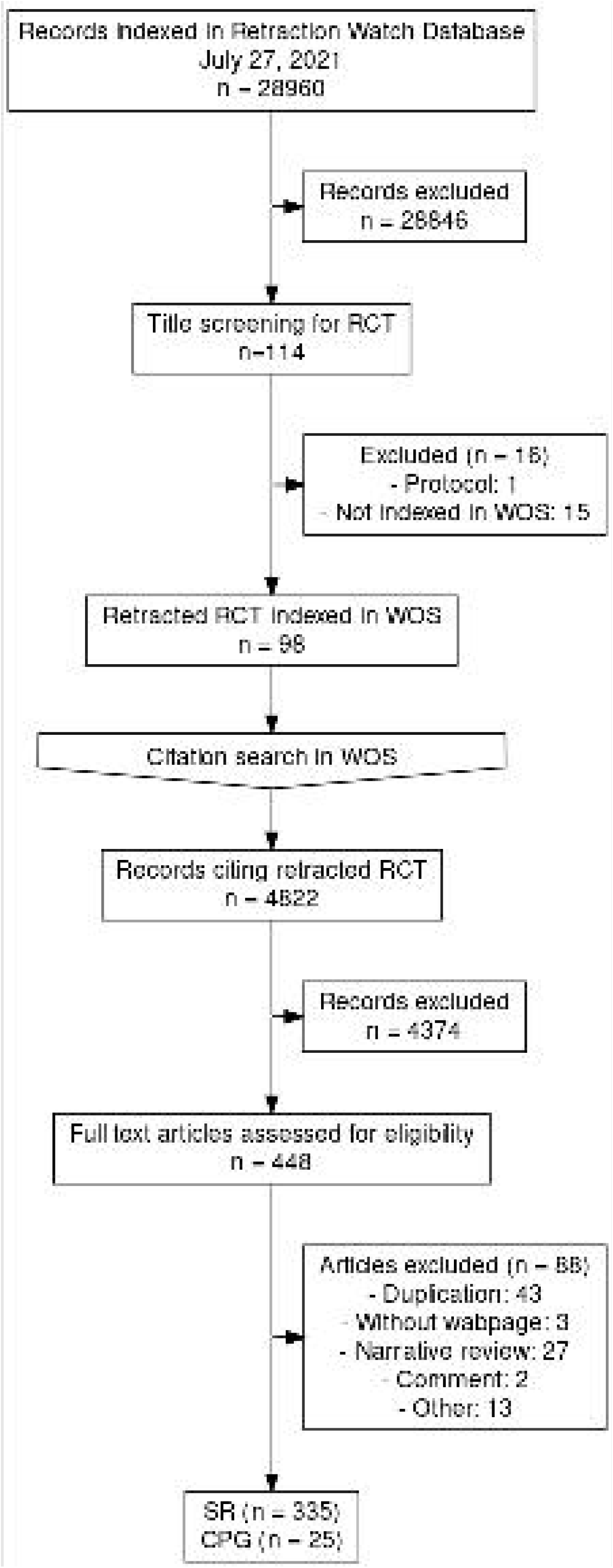
Study flow chart. WOS, Web of Science; RCT, randomised controlled trials; SR, systematic reviews; CPG, clinical practice guidelines

### Characteristics of included articles

Table 1 shows the characteristics of the 98 retracted RCTs. Table S2 summarises their reasons for retraction.

**Table 1.**
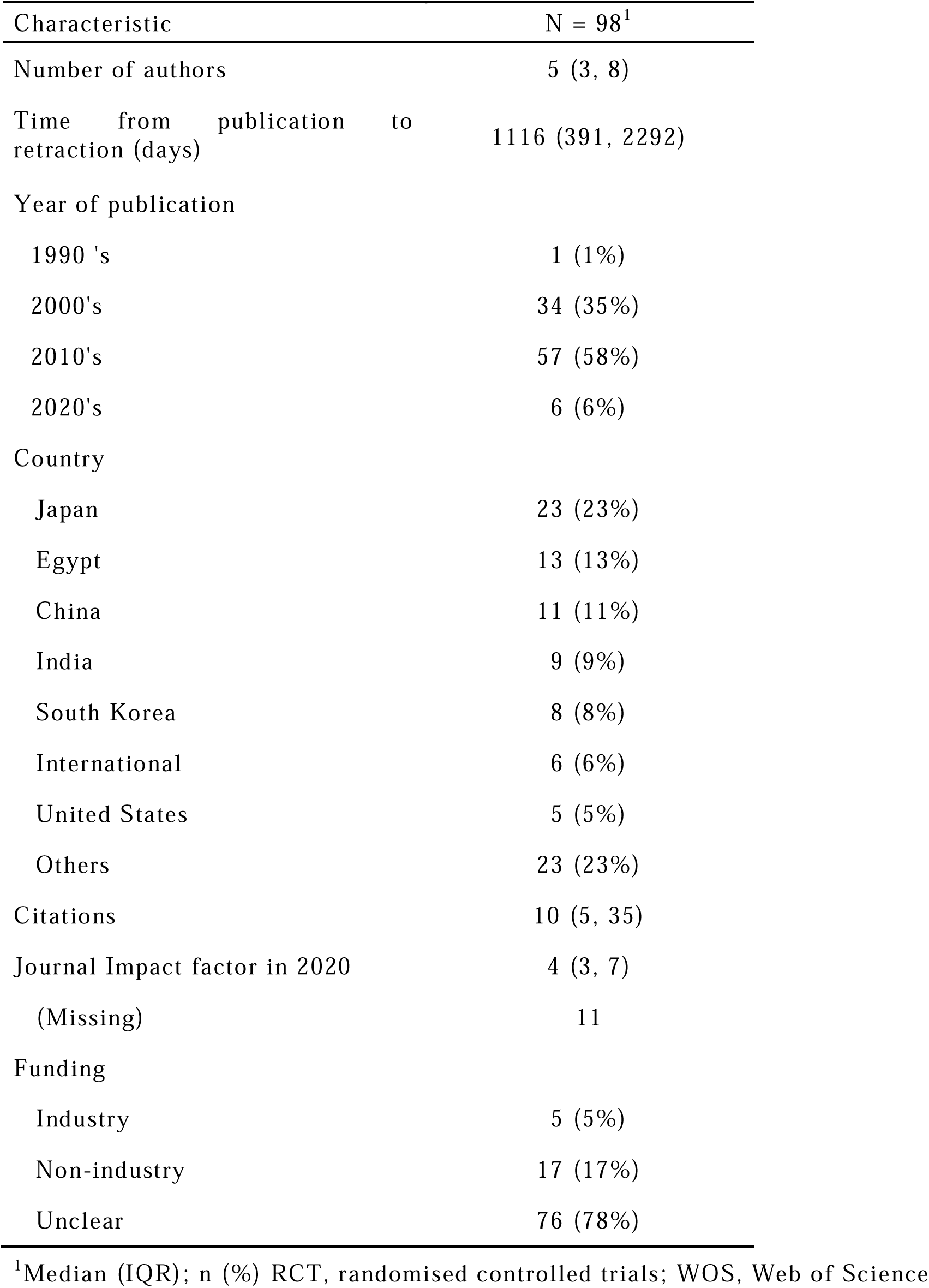
Characteristics of retracted RCTs indexed in WOS.

### Primary outcomes

Of the 335 SRs and 25 CPGs, 157 articles cited RCTs already retracted before their publication, and 203 articles cited RCTs which were later retracted (Tables 2 and 3).

**Table 2.**
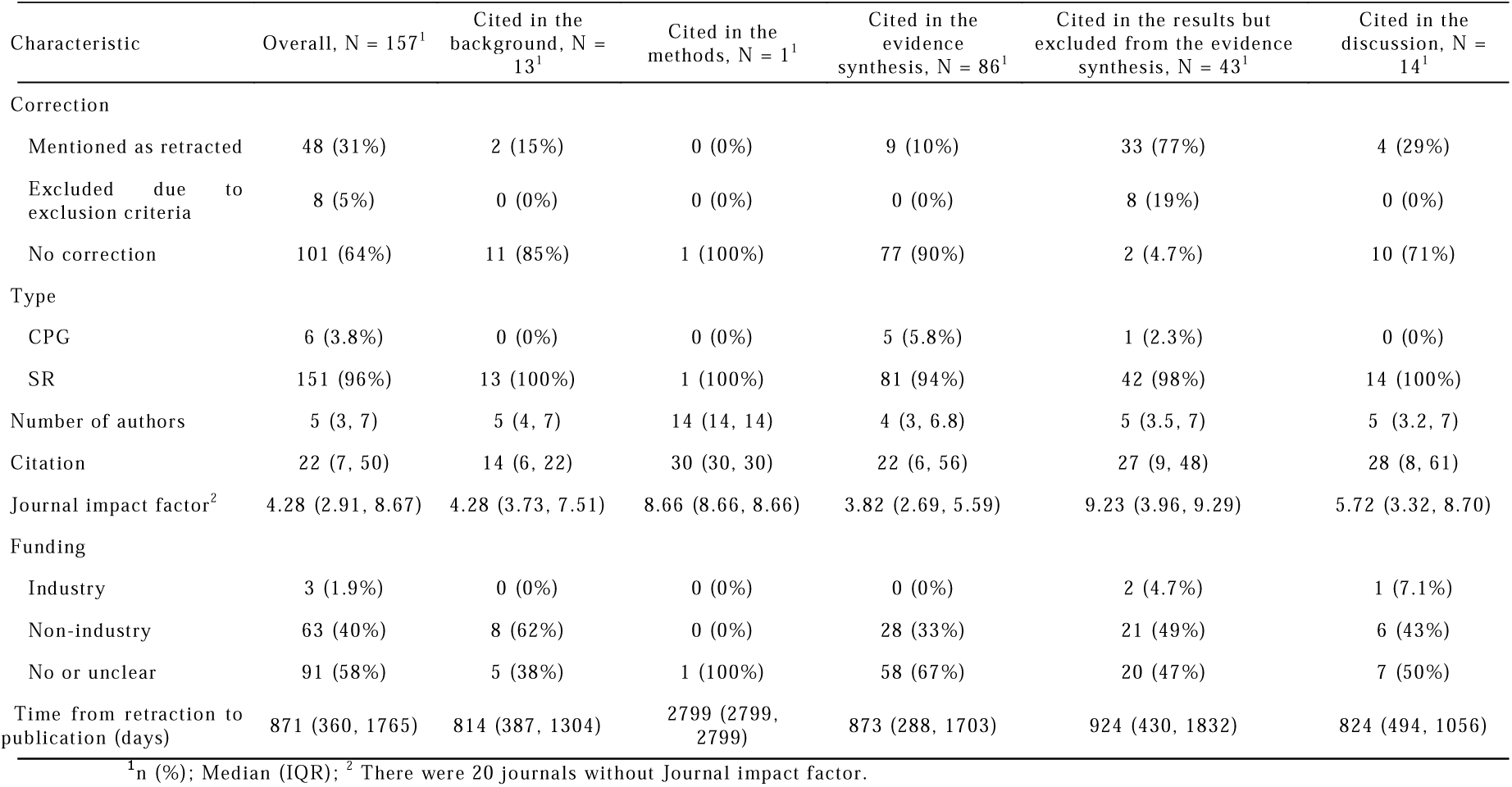
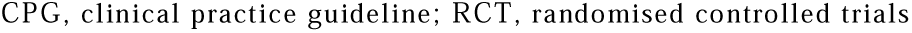
Characteristics of SR and CPG articles which cited already retracted RCTs.

| Characteristic | Overall, N = 157<sup>1</sup> | Cited in the background, N = 13<sup>1</sup> | Cited in the methods, N = 1<sup>1</sup> | Cited in the evidence synthesis, N = 86<sup>1</sup> | Cited in the results but excluded from the evidence synthesis, N = 43<sup>1</sup> | Cited in the discussion, N = 14<sup>1</sup> |
| --- | --- | --- | --- | --- | --- | --- |
| **Correction** |  |  |  |  |  |  |
| Mentioned as retracted | 48 (31%) | 2 (15%) | 0 (0%) | 9 (10%) | 33 (77%) | 4 (29%) |
| Excluded due to exclusion criteria | 8 (5%) | 0 (0%) | 0 (0%) | 0 (0%) | 8 (19%) | 0 (0%) |
| No correction | 101 (64%) | 11 (85%) | 1 (100%) | 77 (90%) | 2 (4.7%) | 10 (71%) |
| **Type** |  |  |  |  |  |  |
| CPG | 6 (3.8%) | 0 (0%) | 0 (0%) | 5 (5.8%) | 1 (2.3%) | 0 (0%) |
| SR | 151 (96%) | 13 (100%) | 1 (100%) | 81 (94%) | 42 (98%) | 14 (100%) |
| Number of authors | 5 (3, 7) | 5 (4, 7) | 14 (14, 14) | 4 (3, 6.8) | 5 (3.5, 7) | 5 (3.2, 7) |
| Citation | 22 (7, 50) | 14 (6, 22) | 30 (30, 30) | 22 (6, 56) | 27 (9, 48) | 28 (8, 61) |
| Journal impact factor<sup>2</sup> | 4.28 (2.91, 8.67) | 4.28 (3.73, 7.51) | 8.66 (8.66, 8.66) | 3.82 (2.69, 5.59) | 9.23 (3.96, 9.29) | 5.72 (3.32, 8.70) |
| **Funding** |  |  |  |  |  |  |
| Industry | 3 (1.9%) | 0 (0%) | 0 (0%) | 0 (0%) | 2 (4.7%) | 1 (7.1%) |
| Non-industry | 63 (40%) | 8 (62%) | 0 (0%) | 28 (33%) | 21 (49%) | 6 (43%) |
| No or unclear | 91 (58%) | 5 (38%) | 1 (100%) | 58 (67%) | 20 (47%) | 7 (50%) |
| Time from retraction to publication (days) | 871 (360, 1765) | 814 (387, 1304) | 2799 (2799, 2799) | 873 (288, 1703) | 924 (430, 1832) | 824 (494, 1056) |

<sup>1</sup>n (%); Median (IQR); <sup>2</sup> There were 20 journals without Journal impact factor.
CPG, clinical practice guideline; RCT, randomised controlled trials

**Table 3.** Characteristics of SR and CPG articles whose cited RCTs were later retracted.

| Characteristic | Overall, N = 203<sup>1</sup> | Cited in the background, N = 14<sup>1</sup> | Cited in the evidence synthesis, N = 149<sup>1</sup> | Cited in the results but excluded, N = 21<sup>1</sup> | Cited in the discussion, N = 19<sup>1</sup> |
| --- | --- | --- | --- | --- | --- |
| Correction |  |  |  |  |  |
| Corrected | 9 (5%)<sup>2</sup> | 0 (0%) | 9 (5%)<sup>2</sup> | 0 (0%) | 0 (0%) |
| Excluded due to the concern about study group | 4 (2%) | 0 (0%) | 0 (0%) | 4 (19%) | 0 (0%) |
| Excluded due to exclusion criteria | 10 (5%) | 0 (0%) | 0 (0%) | 9 (43%) | 0 (0%) |
| No correction | 180 (89%) | 14 (100%) | 140 (95%) | 8 (38%) | 19 (100%) |
| Type |  |  |  |  |  |
| CPG | 19 (9.4%) | 0 (0%) | 18 (12%) | 1 (5%) | 0 (0%) |
| SR | 184 (91%) | 14 (100%) | 131 (88%) | 20 (95%) | 19 (100%) |
| Number of authors | 6 (3, 8) | 4.0 (3, 6) | 6 (5, 7.5) | 5 (3, 9) | 6 (4, 7) |
| Citation | 26 (11, 62) | 26 (6, 61) | 26 (11, 57) | 54 (14, 148) | 38 (16, 74) |
| Journal impact factor<sup>3</sup> | 4.36 (3.31, 8.70) | 4.12 (2.59, 5.60) | 4.17 (3.24, 7.32) | 9.29 (4.46, 9.29) | 5.56 (3.41, 8.78) |
| Funding |  |  |  |  |  |
| Industry | 7 (3.4%) | 1 (7.1%) | 4 (2.7%) | 1 (5%) | 1 (5%) |
| Non-industry | 66 (33%) | 4 (29%) | 41 (28%) | 11 (52%) | 10 (53%) |
| No or unclear | 130 (64%) | 9 (64%) | 104 (70%) | 9 (43%) | 8 (42%) |

<sup>1</sup>n (%); Median (IQR); <sup>2</sup> One SR article was retracted due to plagiarism; <sup>3</sup> There were 20 journals without Journal impact factor. CPG, clinical practice guideline; SR, systematic review

Of the 86 articles citing already retracted RCTs in their evidence synthesis, 77 articles (90%) cited the retracted RCT without caution. The median days from retraction of the included RCT to publication of the citing SR or CPG was 871 days (interquartile range [IQR]: 360, 1765), hence apparently sufficient time to notice the retraction. None of these SRs and CPGs subsequently corrected themselves after their publication up to October 28th, 2021.

A total of 149 articles cited RCTs in their evidence synthesis, which were later retracted. Among them, one SR was retracted due to plagiarism. Results were corrected in 5% of SRs (6/130) and 11% of CPGs (2/18). Figure 2 shows the time from the retraction to correction of these 148 pre-retraction-published articles. The median days from retraction to correction was 962.5 (IQR: 164,1621.5).

**Figure 2.**
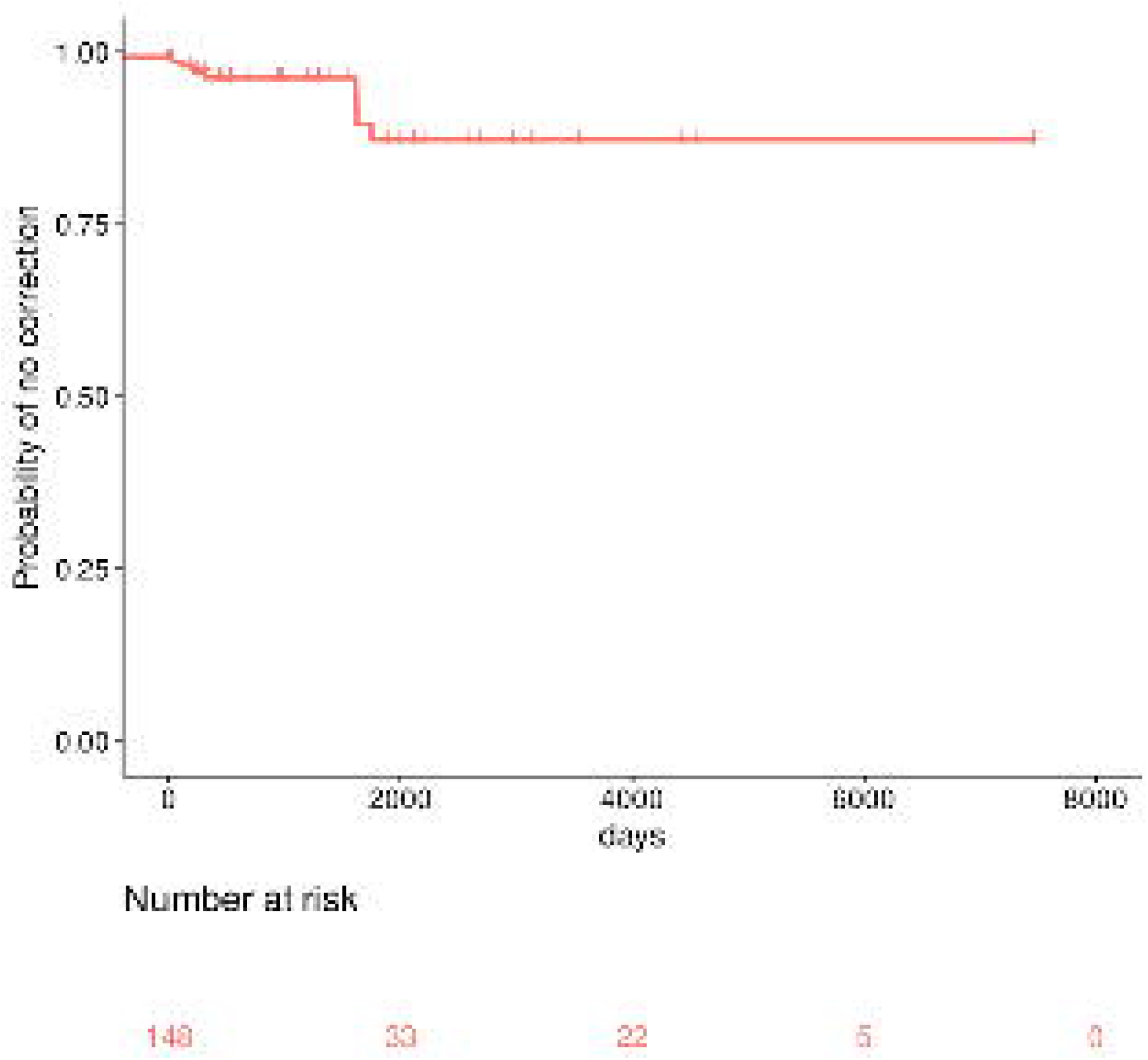
Time from retraction to correction among articles published before retraction.

## Discussion

### Summary of findings

This study presents the most rigorous, comprehensive and up-to-date investigation of the impact of retracted RCTs in the clinically most sensitive literature of SRs and CPGs. We found 77 SRs and CPGs which cited already retracted RCT without cautions, and none of them corrected themselves during a median observation period of more than two years. Of 149 SRs and CPGs which included RCTs that were later retracted in their evidence syntheses, less than one in ten corrected themselves.

### Comparison with other studies

Much attention has been paid recently to the fate and impact of retracted research. Steen evaluated the harmful impact of retracted articles from 2001 to 2010 and found that a large number of patients in trials were treated based on the (mis)information based on retracted articles [19]. Several studies investigated the number of citations after retraction due to scientific misconducts [10,11,20]. Our results indicate that about a half of SRs and CPGs citing already retracted RCTs made inappropriate citations. Avenell et al. investigated the impact of the one osteoporosis research group in doubt. They found that the findings of a third of SRs and CPGs were likely to change if RCTs with concerns were excluded [21]. Hamilton investigated continued citation of retracted radiation oncology articles. He found several SRs and CPGs cited retracted articles without caution [12]. Our results indicate that most inappropriate citations in SRs and CPGs were not corrected regardless of the timing of retractions. Self-correction in scientific community has not improved despite repeated warnings.

### Potential implications

Not to disseminate inappropriate information, both researchers and publishers should be more careful of the possibility of retracted articles.

To decrease the number of SRs and CPGs citing already retracted RCTs, researchers should pay attention to the included articles whether they were retracted. While a previous interrupted time-series analysis has shown that retraction does reduce the number of citations of RCTs [22], the present results indicate that inappropriate citations still prevail. Indeed, retraction is sometimes not reflected in PubMed for a long time [23], and in some cases, it is hard to identify retraction information even in journal web pages [24]. Researchers should check carefully not only the searched abstracts and retrieved full text files but also the journal web pages [25]. Simultaneously, publishers should make the retraction information more clearly discoverable in their web pages. They should also contact databases indexing abstracts such as MEDLINE/ PubMed and repositories such as PubMed Central as soon as possible. MEDLINE/PubMed should increase their efforts to reflect the retraction information in a timely manner. Needless to say, publishers should not publish problematic studies [26].

To improve the timely correction of SRs and CPGs published before retraction, a semi-automated alert system is needed. When an article is retracted, the journal publisher should send the information to publishers citing the retracted article as soon as possible. The Committee on Publication Ethics’ retraction guideline recommend journal editors to ensure retraction information to appear on all online searches for the retracted publication [27]. However, the scope of this recommendation does not include other journals citing retracted articles. This point should be improved.

### Limitations

There are several limitations in our study. First, we searched retracted RCTs in clinical medicine through title searches. If there are studies that did not declare themselves to be randomized trials in their titles, the actual number of SRs and CPGs inappropriately citing retracted RCTs may be greater than the present results. Further study is needed to evaluate the influence of retracted RCTs in a more comprehensive manner. Second, we used the WOS to find citing SRs and CPGs. There is a possibility that we underestimated the impact among those not indexed in the WOS. Third, we did not investigate whether the conclusions and recommendations of SRs and CPGs will change when the retraction RCTs are excluded. Retraction is a concept with a certain range from minor to major [28]. A preliminary analysis including various types of study designs showed that meta-analysis including retracted studies due to issues with data tend to overestimate the effect size [29]. However, we must point out that continuing to cite and to include retracted, hence false, RCTs after their retraction is in itself tantamount to scientific misconduct.

## Conclusions

Many SRs and CPGs cited and included RCTs that had been retracted before their own publication and never corrected themselves since. A great majority of SRs and CPGs that included RCTs which were later retracted did not correct themselves and continued to be available even after one or more of their included RCTs were retracted. The whole scientific and medical community including publishers and researchers should make efforts to remove the impact of retracted RCTs.

## Data Availability

Dataset available from the corresponding author upon the request.

## Acknowledgements

Authors wish to thank Dr. Akihiro Shiroshita for retrieving articles.

## Footnotes

### Contributors

YK had full access to all the data in the study and take responsibility for the integrity of the data and the accuracy of the data analysis. Study concept and design: YK, MB,

YT, and TAF. Acquisition of data: YK, MB, YT, TA, ST, TS, and SO. Analysis and interpretation of data: YK, MB, YT, TA, ST, TS, SO, and TAF. Drafting of the manuscript: YK. Critical revision of the manuscript for important intellectual content: MB, YT, TA, ST, TS, SO, and TAF. All authors gave final approval of the version to be published and agreed to be accountable for all aspects of this work.

### Ethical approval

Not required.

### Data sharing

Dataset available from the corresponding author upon the request.

### Transparency statement

The lead author affirms that this manuscript is an honest, accurate, and transparent account of the study being reported; that no important aspects of the study have been omitted; and that any discrepancies from the study as planned have been explained.

### Dissemination to participants and related patient and public communities

We plan to present our findings at a national scientific meeting. We also plan to use social media outlets to disseminate findings.

