## supplement for "The impact of retracted randomised controlled trials on systematic reviews and clinical practice guidelines: a meta-epidemiological study"

Table S1. Items for reporting methodological research, adapted from the PRISMA Checklist

| Section/topic | # | | Checklist item | Reported on page # |
| --- | --- | --- | --- | --- |
| TITLE | | | |  |
| Title | 1 | | Identify the report as a meta-epidemiologic study. | 1 |
| ABSTRACT | | | |  |
| Structured summary | 2 | | Provide a structured summary that includes the background of the topic, goal of the study, data sources, method of data selection, appraisal and synthesis methods, results, limitations, conclusions and implications of key findings. | 3-4 |
| INTRODUCTION | | | |  |
| Rationale | 3 | | Describe the rationale for the meta-epidemiological study in the context of what is already known. | 6 |
| Objectives | 4 | | Provide an explicit statement of the goal of the meta-epidemiological study and the hypothesis being empirically tested. | 6 |
| METHODS | | | |  |
| Protocol | 5 | | Indicate if a protocol exists, if and where it can be accessed (eg, Web address). Registration of a protocol is not mandatory | 6 |
| Eligibility criteria | 6 | | Specify study characteristics used as criteria for eligibility with a rationale. | 7 |
| Information sources | 7 | | Describe all information sources (eg, databases with dates of coverage, contact with experts to identify additional studies, Internet searches) and search date. | 7 |
| Search | 8 | | Present full electronic search strategy for at least one database, including any limits used, such that it could be repeated. Search is commonly not driven by a clinical question. | 7 |
| Study selection | 9 | | Describe the process for selecting studies for inclusion (ie, how many reviewers selected studies, reviewing in duplicate or by single individuals). | 7 |
| Data collection process | 10 | | Describe method of data extraction from reports (eg, piloted forms, independently, in duplicate) and any processes used for manipulating data or obtaining and confirming data from investigators. | 8 |
| Data items | 11 | | List and define all variables for which data were sought and any assumptions and imputations made. | 8-9 |
| Risk of bias in individual studies | 12 | | If risk of bias assessment of individual studies was relevant to the analysis, describe the items used and how this information is to be used during data synthesis. | NA |
| Summary measures | 13 | | State the principal summary measures (eg, ratio of risk ratios, difference in means) and explain its meaning and direction to readers. | 8 |
| Synthesis of results | 14 | | Describe the statistical or descriptive methods of synthesis including measures of consistency if relevant. If applicable, describe the development of statistical or simulation modelling based on theoretical background. Describe and justify assumptions and computational approximations. Describe methods of additional analyses (eg, sensitivity or subgroup analyses, meta-regression), if done, indicating which were prespecified. | 9 |
| RESULTS | | | |  |
| Study selection | | 15 | Give numbers of studies assessed for eligibility and included in the study, with reasons for exclusions at each stage, ideally with a flow diagram. Present a measure of inter-reviewer agreement (eg, kappa statistic). | 9-10 |
| Study characteristics | | 16 | For each study, present characteristics for which data were extracted and provide the citations. Clinical characteristics may not always be relevant. | 10 |
| Risk of bias within studies | | 17 | If risk of bias assessment of individual studies was used in the meta-epidemiological analysis, report risk of bias indicators of each study to allow replication of findings. | NA |
| Results of individual studies | | 18 | Present data elements used in the meta-epidemiological analysis from each study (results of clinical outcomes may not be relevant). | NA |
| Synthesis of results | | 19 | Present results of statistical analysis done, including measures of precision and measures of consistency. Present validity of assumptions and fit of statistical or simulation modelling, if applicable. | 10 |
| Additional analysis | | 20 | Give results of additional analyses, if done (eg, sensitivity or subgroup analyses, meta-regression). | 10 |
| DISCUSSION | | | |  |
| Summary of evidence | | 21 | Summarise the main findings and compare them with existing knowledge about the topic. The quality of evidence may not be relevant; however, investigators should describe their certainty in the results to readers. | 11 |
| Limitations | | 22 | Discuss limitations at research methodology level (eg, likelihood of reporting or publication bias). | 12-13 |
| Conclusions | | 23 | Provide general interpretation of the results and implications for future research. Provide any plausible impact on clinical practice. | 13 |
| FUNDING | | | |  |
| Funding | | 24 | Describe sources of funding for the methodology research and role of funders. | 14 |

*From:* Murad MH, Wang Z. Guidelines for reporting meta-epidemiological methodology research. Evid Based Med. 2017 Aug;22(4):139–42.

Table S2. Retracted randomised controlled trials

| **Title** | **Journal** | **Publisher** | **OriginalPaperDOI** | **OriginalWOSID** |
| --- | --- | --- | --- | --- |
| Negative pressure wound therapy versus conventional dressing for open fractures in lower extremity trauma: a multicentre randomized controlled trial | The Bone and Joint Journal | Bone & Joint Publishing: The British Editorial Society of Bone & Joint Surgery | 10.1302/0301-620X.102B7.BJJ-2019-1462.R1 | WOS:000561874100016 |
| A training programme involving automatic self-transcending meditation in late-life depression: preliminary analysis of an ongoing randomised controlled trial | BJPsych Open | Cambridge University Press | 10.1192/bjpo.bp.115.002394 | WOS:000408510800015 |
| Effects of vitamin D supplementation on glucose metabolism, lipid concentrations, inflammation, and oxidative stress in gestational diabetes: a double-blind randomized controlled clinical trial | The American Journal of Clinical Nutrition | Oxford Academic | 10.3945/ajcn.113.072785 | WOS:000328002000009 |
| Magnesium supplementation affects metabolic status and pregnancy outcomes in gestational diabetes: a randomized, double-blind, placebo-controlled trial | The American Journal of Clinical Nutrition | Oxford Academic | 10.3945/ajcn.114.098616 | WOS:000357425500030 |
| Decompressive surgery in patients with malignant middle cerebral artery infarction: A randomized, controlled trial in a Turkish population (Demitur trial) | International Journal of Stroke | SAGE Publications | 10.1177/17474930211007671 | WOS:000650030400001 |
| Purified palmitoleic acid for the reduction of high-sensitivity C-reactive protein and serum lipids: a double-blinded, randomized, placebo controlled study | Journal of Clinical Lipidology | Elsevier | 10.1016/j.jacl.2014.08.001 | WOS:000662981800018 |
| Skin rejuvenation by microneedle fractional radiofrequency and a human stem cell conditioned medium in Asian skin: a randomized controlled investigator blinded split-face study | Journal of Cosmetic and Laser Therapy: Official Publication of the European Society for Laser Dermatology | Taylor and Francis | 10.3109/14764172.2012.748201 | WOS:000314306600005 |
| ORIENT-3: A randomized, open-label, phase III study of sintilimab versus docetaxel in previously treated advanced/metastatic squamous non-small cell lung cancer (sqNSCLC) | Annals of Oncology : Official Journal of the European Society for Medical Oncology ESMO | Elsevier | 10.1016/j.annonc.2020.10.517 | WOS:000600992500030 |
| Efficacy and safety of tripterygium glycosides for active moderate to severe Graves' ophthalmopathy: a randomised, observer-masked, single-centre trial | European journal of Endocrinology (European Federation of Endocrine Societies) | BioScientifica | 10.1530/EJE-20-0857 | WOS:000608421100013 |
| Long-term follow-up after sleeve gastrectomy versus Roux-en-Y gastric bypass versus one-anastomosis gastric bypass: a prospective randomized comparative study of weight loss and remission of comorbidities | Surgical Endoscopy | Springer | 10.1007/s00464-018-6307-9 | WOS:000456976700006 |
| Successful implementation of an enhanced recovery after surgery (ERAS) protocol reduces nausea and vomiting after infratentorial craniotomy for tumour resection: a randomized controlled trial | BMC Neurology | Springer - Biomed Central (BMC) | 10.1186/s12883-020-01699-z | WOS:000530142700002 |
| Bismuth adjuvant ameliorates adverse effects of high-dose chemotherapy in patients with multiple myeloma and malignant lymphoma undergoing autologous stem cell transplantation: a randomised, double-blind, prospective pilot study | Supportive Care in Cancer | Springer | 10.1007/s00520-016-3522-6 | WOS:000394991500033 |
| Anastrozole or letrozole for ovulation induction in clomiphene-resistant women with polycystic ovarian syndrome: a prospective randomized trial | Fertility and Sterility | Elsevier | 10.1016/j.fertnstert.2007.05.010 | WOS:000256075700025 |
| Clomiphene citrate or aromatase inhibitors for superovulation in women with unexplained infertility undergoing intrauterine insemination: a prospective randomized trial | Fertility and Sterility | Elsevier | 10.1016/j.fertnstert.2008.06.013 | WOS:000270616100037 |
| Clomiphene citrate or letrozole for ovulation induction in women with polycystic ovarian syndrome: a prospective randomized trial | Fertility and Sterility | Elsevier | 10.1016/j.fertnstert.2007.02.062 | WOS:000269711700002 |
| Letrozole versus combined metformin and clomiphene citrate for ovulation induction in clomiphene-resistant women with polycystic ovary syndrome: a randomized controlled trial | Fertility and Sterility | Elsevier | 10.1016/j.fertnstert.2009.07.985 | WOS:000281674600040 |
| Artemisia annua and Artemisia afra tea infusions vs. artesunate-amodiaquine (ASAQ) in treating Plasmodium falciparum malaria in a large scale, double blind, randomized clinical trial | Phytomedicine | Elsevier | 10.1016/j.phymed.2018.12.002 | WOS:000465081700006 |
| A randomized comparison of ultrasound-guided versus landmark-based corticosteroid injection for trigger finger | Journal of Hand Surgery (European Volume) | SAGE Publications | 10.1177/1753193419839892 | WOS:000559528700001 |
| Ultrasound-guided versus blind corticosteroid injections for De Quervain tendinopathy: a prospective randomized trial | Journal of Hand Surgery (European Volume) | SAGE Publications | 10.1177/1753193418790535 | WOS:000444583100005 |
| Comparison of 0.05% cyclosporine and 3% diquafosol solution for dry eye patients: a randomized, blinded, multicenter clinical trial | BMC Ophthalmology | Springer - Biomed Central (BMC) | 10.1186/s12886-019-1136-8 | WOS:000471892300001 |
| Greater analgesic effect with intermittent compared with continuous mode of lumbar plexus block for total hip arthroplasty: a randomized controlled trial | Regional Anesthesia and Pain Medicine | BMJ Publishing | 10.1136/rapm-2018-100091 | WOS:000471158500005 |
| Comparison of sugammadex and pyridostigmine bromide for reversal of rocuronium-induced neuromuscular blockade in short-term pediatric surgery: A prospective randomized study | Medicine | Wolters Kluwer - Lippincott Williams & Wilkins | 10.1097/MD.0000000000019130 | WOS:000525861300057 |
| A comparison between intrathecal clonidine and neostigmine as an adjuvant to bupivacaine in the subarachnoid block for elective abdominal hysterectomy operations: A prospective, double-blind and randomized controlled study | Saudi Journal of Anaesthesia | Wolters Kluwer - Medknow | 10.4103/1658-354X.168797 | WOS:000386755200002 |
| Comparative evaluation of analgesic sparing efficacy between dexmedetomidine and clonidine used as adjuvant to ropivacaine in thoracic paravertebral block for patients undergoing breast cancer surgery: A prospective, randomized, double-blind study | Saudi Journal of Anaesthesia | Wolters Kluwer - Medknow | 10.4103/sja.SJA_81_18 | WOS:000446489400010 |
| Controlled hypotension in day care functional endoscopic sinus surgery: A comparison between esmolol and dexmedetomidine: A prospective, double-blind, and randomized study | Saudi Journal of Anaesthesia | Wolters Kluwer - Medknow | 10.4103/1658-354X.174919 | WOS:000386755600007 |
| Effect of dexmedetomidine as adjuvant in ropivacaine-induced supraclavicular brachial plexus block: A prospective, double-blinded and randomized controlled study | Saudi Journal of Anaesthesia | Wolters Kluwer - Medknow | 10.4103/1658-354X.144082 | WOS:000219143500016 |
| Pain relief after ambulatory hand surgery: A comparison between dexmedetomidine and clonidine as adjuvant in axillary brachial plexus block: A prospective, double-blinded, randomized controlled study | Saudi Journal of Anaesthesia | Wolters Kluwer - Medknow | 10.4103/1658-354X.169443 | WOS:000386754400003 |
| Pain relief after Arthroscopic Knee Surgery: A comparison of intra-articular ropivacaine, fentanyl, and dexmedetomidine: A prospective, double-blinded, randomized controlled study | Saudi Journal of Anaesthesia | Wolters Kluwer - Medknow | 10.4103/1658-354X.130727 | WOS:000219136100017 |
| Pain relief in day care arthroscopic knee surgery: A comparison between intra-articular ropivacaine and levobupivacaine: A prospective, double-blinded, randomized controlled study | Saudi Journal of Anaesthesia | Wolters Kluwer - Medknow | 10.4103/1658-354X.136435 | WOS:000219137500013 |
| PONV in Ambulatory surgery: A comparison between Ramosetron and Ondansetron: a prospective, double-blinded, and randomized controlled study | Saudi Journal of Anaesthesia | Wolters Kluwer - Medknow | 10.4103/1658-354X.125917 | WOS:000219132400007 |
| Cervical mucus removal prior to intrauterine insemination: a randomized trial | BJOG: British Journal of Obstetrics and Gynaecology | Wiley | 10.1111/1471-0528.15003 | WOS:000433566700014 |
| A parallel randomized controlled trial examining the effects of rhythmic sensory stimulation on fibromyalgia symptoms | PLoS One | PLoS | 10.1371/journal.pone.0212021 | WOS:000460371600006 |
| Obturator nerve block transurethral surgery for bladder cancer: comparison of inguinal and intravesical approaches: prospective randomized trial | Irish Journal of Medical Science | Springer | 10.1007/s11845-015-1300-y | WOS:000379758700002 |
| Peri-conceptional progesterone treatment in women with unexplained recurrent miscarriage: a randomized double-blind placebo-controlled trial | The Journal of Maternal-Fetal Medicine | Taylor and Francis | 10.1080/14767058.2017.1286315 | WOS:000423819700021 |
| Calcium versus oral contraceptive pills containing drospirenone for the treatment of mild to moderate premenstrual syndrome: A double blind randomized placebo controlled trial | European Journal of Obstetrics, Gynecology, and Reproductive Biology | Elsevier | 10.1016/j.ejogrb.2016.01.015 | WOS:000371836500019 |
| Effect of a high-fat Mediterranean diet on bodyweight and waist circumference: a prespecified secondary outcomes analysis of the PREDIMED randomised controlled trial | The Lancet: Diabetes & Endocrinology | Elsevier | 10.1016/S2213-8587(16)30085-7 | WOS:000380761600020 |
| Denosumab versus zoledronic acid in cases of surgically unsalvageable giant cell tumor of bone: A randomized clinical trial | Journal of Bone Oncology | Elsevier | 10.1016/j.jbo.2019.100217 | WOS:000468770500008 |
| Dexmedetomidine in a surgically inserted catheter for transversus abdominis plane block in donor hepatectomy: A prospective randomized controlled study | Saudi Journal of Anaesthesia | Wolters Kluwer | 10.4103/sja.SJA_577_17 | WOS:000446923200022 |
| Digital assistance of nasogastric tube insertion in intubated patients under general anesthesia: A single-blinded prospective randomized study | Saudi Journal of Anaesthesia | Wolters Kluwer | 10.4103/sja.SJA_524_16 | WOS:000404816800005 |
| Cardiac stem cells in patients with ischaemic cardiomyopathy (SCIPIO): initial results of a randomised phase 1 trial | Lancet | Elsevier | 10.1016/S0140-6736(11)61590-0 | WOS:000297695800029 |
| Effect of perioperative infusion of Dexmedetomidine combined with Sufentanil on quality of postoperative analgesia in patients undergoing laparoscopic nephrectomy: a CONSORT-prospective, randomized, controlled trial | BMC Anesthesiology | Springer - Biomed Central (BMC) | 10.1186/s12871-018-0608-3 | WOS:000447846300001 |
| The effectiveness of the Invisalign appliance in extraction cases using the the ABO model grading system: a multicenter randomized controlled trial | International Journal of Clinical and Experimental Medicine | e-Century Publishing Corporation | unavailable | WOS:000359293200210 |
| Reduction in the incidence of type 2 diabetes with the Mediterranean diet: results of the PREDIMED-Reus nutrition intervention randomized trial | Diabetes Care | American Diabetes Association | 10.2337/dc10-1288 | WOS:000286497000004 |
| A comparison of granisetron, droperidol, and metoclopramide in the treatment of established nausea and vomiting after breast surgery: A double-blind, randomized, controlled trial | Clinical Therapeutics | Elsevier | 10.1016/S0149-2918(03)80072-3 | WOS:000182628200007 |
| Effects of dexamethasone in preventing postoperative emetic symptoms after total knee replacement surgery: A prospective, randomized, double-blind, vehicle-controlled trial in adult Japanese patients | Clinical Therapeutics | Elsevier | 10.1016/j.clinthera.2005.05.011 | WOS:000230531000007 |
| Flurbiprofen axetil preceded by venous occlusion in the prevention of pain on propofol injection in the hand: A prospective, randomized, double-blind, vehicle-controlled, dose-finding study in Japanese adult surgical patients | Clinical Therapeutics | Elsevier | 10.1016/j.clinthera.2005.05.003 | WOS:000229558700007 |
| Granisetron versus granisetron/dexamethasone combination for the treatment of nausea, retching, and vomiting after major gynecologic surgery: A randomized, double-blind study | Clinical Therapeutics | Elsevier | 10.1016/S0149-2918(03)80092-9 | WOS:000181433900009 |
| Influence of age on flurbiprofen axetil requirements for preventing pain on injection of propofol in japanese adult surgical patients: A prospective, randomized, double-blind, vehicle-controlled, parallel-group, dose-ranging study | Clinical Therapeutics | Elsevier | 10.1016/j.clinthera.2006.08.015 | WOS:000240521000004 |
| Prevention of pain due to injection of propofol with IV administration of lidocaine 40 mg + metoclopramide 2.5, 5, or 10 mg or saline: A randomized, double-blind study in Japanese adult surgical patients | Clinical Therapeutics | Elsevier | 10.1016/j.clinthera.2007.05.019 | WOS:000247335600007 |
| Results of a prospective, randomized, double-blind, placebo-controlled, dose-ranging trial to determine the effective dose of ramosetron for the prevention of vomiting after tonsillectomy in children | Clinical Therapeutics | Elsevier | 10.1016/S0149-2918(03)90097-X | WOS:000188089600009 |
| Treatment of postoperative emetic symptoms with granisetron in women undergoing abdominal hysterectomy: a randomized, double-blind, placebo-controlled, dose-ranging study | Current Therapeutic Research | Elsevier | 10.1016/j.curtheres.2004.06.002 | WOS:000223707800001 |
| Comparison of the effects of extracorporeal shock wave therapy and a vacuum erectile device on penile erectile dysfunction: a randomized clinical trial | Medicine | Wolters Kluwer | 10.1097/MD.0000000000008414 | WOS:000415106800025 |
| Effect of acupuncture and Chinese herbal medicine on subacute stroke outcomes: a single center randomized controlled trial | Acupuncture in Medicine | BMJ Publishing | 10.1136/acupmed-2016-011167 | WOS:000448518900011 |
| High-concentration supplemental perioperative oxygen and surgical site infection following elective colorectal surgery for rectal cancer: A prospective, randomized, double-blind, controlled, single-site trial | American Journal of Surgery | Elsevier | 10.1016/j.amjsurg.2014.04.002 | WOS:000344935500006 |
| The effect of postural control intervention for congenital muscular torticollis: a randomized controlled trial | Clinical Rehabilitation | SAGE Publications | 10.1177/0269215514555037 | WOS:000358880000007 |
| Etidronate for fracture prevention in amyotrophic lateral sclerosis: A randomized controlled trial | Bone | Elsevier | 10.1016/j.bone.2006.04.025 | WOS:000241584500016 |
| The effect of Polygonum minus extract on cognitive and psychosocial parameters according to mood status among middle-aged women: a randomized, double-blind, placebo-controlled study | Clinical Interventions in Aging | Taylor and Francis - Dove Press | 10.2147/CIA.S86411 | WOS:000361614100001 |
| Once-weekly risedronate for prevention of hip fracture in women with Parkinson's disease: a randomised controlled trial | Journal of Neurology, Neurosurgery, and Psychiatry | BMJ Publishing | 10.1136/jnnp.2011.244574 | WOS:000296766100019 |
| Intracytoplasmic morphologically selected sperm injection versus conventional intracytoplasmic sperm injection: a randomized controlled trial | Reproductive Biology and Endocrinology (RB&E) | BioMed Central (BMC) | 10.1186/s12958-015-0096-y | WOS:000360278700001 |
| Evaluation of the risk of postcesarean endometritis with preoperative vaginal preparation with povidoneâ€“iodine: A randomized controlled study | Middle East Fertility Society Journal | Elsevier | 10.1016/j.mefs.2015.03.002 | WOS:000365187300006 |
| A randomized controlled trial of botulinum toxin A for treating neuropathic pain in patients with spinal cord injury | Medicine | Wolters Kluwer - Lippincott Williams & Wilkins | 10.1097/MD.0000000000006919 | WOS:000401821600026 |
| Comparison of four facial muscles, orbicularis oculi, corrugator supercilii, masseter or mylohyoid, as best predictor of good conditions for intubation: A randomised blinded trial. | European Journal of Anaesthesiology | Wolters Kluwer | 10.1097/EJA.0b013e3283625039 | WOS:000326601800007 |
| The prevention of hip fracture with risedronate and ergocalciferol plus calcium supplementation in elderly women with Alzheimer disease: A randomized controlled trial | Archives of Internal Medicine | JAMA Network | 10.1001/archinte.165.15.1737 | WOS:000231034800010 |
| Postoperative pain after irrigation with Vibringe versus a conventional needle: a randomized controlled trial | International Endodontic Journal | Wiley | 10.1111/iej.12615 | WOS:000380036400012 |
| Effect of folate and mecobalamin on hip fractures in patients with stroke: a randomized controlled trial | JAMA: Journal of the American Medical Association | American Medical Association | 10.1001/jama.293.9.1082 | WOS:000227285200018 |
| Prospective, randomized, controlled trial of silicate-substituted calcium phosphate versus rhBMP-2 in a minimally invasive transforaminal lumbar interbody fusion | Spine | Lippincott Williams and Wilkins | 10.1097/BRS.0000000000000106 | WOS:000336211800015 |
| Effect of nitroglycerin ointment on bone density and strength in postmenopausal women: a randomized trial | JAMA: Journal of the American Medical Association | JAMA Network | 10.1001/jama.2011.176 | WOS:000287594300023 |
| A phase III, randomized, double-blind, matched-pairs, active-controlled clinical trial and preclinical animal study to compare the durability, efficacy and safety between polynucleotide filler and hyaluronic acid filler in the correction of crow's feet: a new concept of regenerative filler | Journal of Korean Medical Science | The Korean Academy of Medical Sciences | 10.3346/jkms.2014.29.S3.S201 | WOS:000354631700008 |
| Effect of ramipril on walking times and quality of life among patients with peripheral artery disease and intermittent claudication: a randomized controlled trial. | JAMA: Journal of the American Medical Association | American Medical Association | 10.1001/jama.2012.216237 | WOS:000314466900026 |
| Total knee arthroplasty performed with either a mini-subvastus or a standard approach: a prospective randomized controlled study with a minimum follow-up of 2 years | Archives of Orthopedic and Trauma Surgery | Springer - Nature Publishing Group | 10.1007/s00402-014-1963-2 | WOS:000339871100014 |
| Lamotrigine in the immediate treatment of outpatients with depersonalization disorder without psychiatric comorbidity: randomized, double-blind, placebo-controlled study | Journal of Clinical Psychopharmacology | Wolters Kluwer - Lippincott Williams & Wilkins | 10.1097/JCP.0b013e31820428e1 | WOS:000285771000011 |
| A randomized, double-blind, placebo-controlled study assessing the anti-inflammatory effects of ketamine in cardiac surgical patients | Journal of Cardiothoracic and Vascular Anesthesia | Elsevier | 10.1053/j.jvca.2005.12.005 | WOS:000237072000014 |
| Manuka honey vs. hydrogel--a prospective, open label, multicentre, randomised controlled trial to compare desloughing efficacy and healing outcomes in venous ulcers | Journal of Clinical Nursing | Wiley | 10.1111/j.1365-2702.2008.02558.x | WOS:000262476100018 |
| Efficacy of granisetron for the treatment of postoperative nausea and vomiting in women undergoing breast surgery: a randomised, double-blind, placebo-controlled trial | Clinical Drug Investigation | Springer | 10.2165/00044011-200626040-00004 | WOS:000237687200004 |
| Transanal haemorrhoidal dearterialisation with mucopexy versus stapler haemorrhoidopexy: a randomised trial with long-term follow-up | Annals of the Royal College of Surgeons of England | Royal Society Publishing | 10.1308/003588413X13511609958136 | WOS:000318827600003 |
| Physical activity within a CBT intervention improves coping with pain in traumatized refugees: results of a randomized controlled design. | Pain Medicine: The Official Journal of the American Academy of Pain Medicine | Oxford Academic | 10.1111/j.1526-4637.2010.01040.x | WOS:000287200000008 |
| Prevention of postoperative nausea and vomiting with a small dose of propofol alone and combined with dexamethasone in patients undergoing laparoscopic cholecystectomy: a prospective, randomized, double-blind study | Surgical Endoscopy | Springer | 10.1007/s00464-007-9647-4 | WOS:000255999700023 |
| Effects of a pre-visit educational website on information recall and needs fulfilment in breast cancer genetic counselling, a randomized controlled trial | Breast Cancer Research: BCR | BioMed Central (BMC) | 10.1186/bcr3133 | WOS:000304771800012 |
| A randomized clinical study of circumcision with a ring device versus conventional circumcision | The Journal of Urology | Elsevier | 10.1016/j.juro.2012.07.048 | WOS:000310438600063 |
| Outcome of short proximal femoral nail antirotation and dynamic hip screw for fixation of unstable trochanteric fractures. A randomised prospective comparative trial | Hip International: the Journal of Clinical and Experimental Research on Hip Pathology and Therapy | Wichtig Publishing | 10.5301/HIP.2011.8657 | WOS:000295973300004 |
| Omega-3 fatty acids, vitamin C and Zn supplementation in asthmatic children: a randomized self-controlled study | Acta Paediatrica | Wiley | 10.1111/j.1651-2227.2008.01213.x | WOS:000263965400030 |
| Prophylaxis with oral granisetron for the prevention of nausea and vomiting after laparoscopic cholecystectomy: a prospective randomized study | Archives of Surgery | American Medical Association | 10.1001/archsurg.136.1.101 | WOS:000166307500023 |
| Pretreatment with flurbiprofen axetil, flurbiprofen axetil preceded by venous occlusion, and a mixture of flurbiprofen axetil and propofol in reducing pain on injection of propofol in adult Japanese surgical patients: A prospective, randomized, double-blind, placebo-controlled study | Clinical Therapeutics | Elsevier | 10.1016/j.clinthera.2009.04.014 | WOS:000266182900004 |
| A comparison of pretreatment with fentanyl and lidocaine preceded by venous occlusion for reducing pain on injection of propofol: A prospective, randomized, double-blind, placebo-controlled study in adult Japanese surgical patients | Clinical Therapeutics | Elsevier | 10.1016/j.clinthera.2009.10.012 | WOS:000272018300005 |
| A prospective, randomized, double-blind, placebo-controlled study to assess the antiemetic effects of midazolam on postoperative nausea and vomiting in women undergoing laparoscopic gynecologic surgery | Clinical Therapeutics | Elsevier | 10.1016/j.clinthera.2010.08.005 | WOS:000281688800006 |
| Comparison of lidocaine, metoclopramide, and flurbiprofen axetil for reducing pain on injection of propofol in Japanese adult surgical patients: A prospective, randomized, double-blind, parallel-group, placebo-controlled study | Clinical Therapeutics | Elsevier | 10.1016/j.clinthera.2008.02.018 | WOS:000254466900006 |
| Comparison of propofol, droperidol, and metoclopramide for prophylaxis of postoperative nausea and vomiting after breast cancer surgery: A prospective, randomized, double-blind, placebo-controlled study in Japanese patients | Clinical Therapeutics | Elsevier | 10.1016/j.clinthera.2008.11.011 | WOS:000261811600007 |
| Long-term survival results of a randomized trial comparing gemcitabine/cisplatin and methotrexate/vinblastine/doxorubicin/cisplatin in patients with locally advanced and metastatic bladder cancer | Annals of Oncology : Official Journal of the European Society for Medical Oncology ESMO | Oxford University Press | 10.1093/annonc/mdj965 | WOS:000296292700034 |
| Submucous myomas and their implications in the pregnancy rates of patients with otherwise unexplained primary infertility undergoing hysteroscopic myomectomy: a randomized matched control study | Fertility and Sterility | Elsevier | 10.1016/j.fertnstert.2009.03.075 | WOS:000279758800048 |
| Safety and efficacy of sildenafil citrate in the treatment of Parkinson-emergent erectile dysfunction: a double-blind, Placebo-controlled, randomized study | International Journal of Impotence Research | Springer - Nature Publishing Group | 10.1038/ijir.2010.23 | WOS:000283165100005 |
| Combination treatment of angiotensin-II receptor blocker and angiotensin-converting-enzyme inhibitor in non-diabetic renal disease (COOPERATE): a randomised controlled trial | Lancet | Elsevier | 10.1016/S0140-6736(03)12229-5 | WOS:000180428000010 |
| Single autologous stem-cell transplantation followed by maintenance therapy with thalidomide is superior to double autologous transplantation in multiple myeloma: results of a multicenter randomized clinical trial | Blood | American Society of Hematology | 10.1182/blood-2007-07-101212 | WOS:000253251100021 |
| Chemoembolization combined with radiofrequency ablation for patients with hepatocellular carcinoma larger than 3 cm: a randomized controlled trial. | JAMA: Journal of the American Medical Association | American Medical Association | 10.1001/jama.299.14.1669 | WOS:000254749600023 |
| A prospective randomized trial on the role of perioperative celecoxib administration for total knee arthroplasty: improving clinical outcomes | Anesthesia & Analgesia | Wolters Kluwer - Lippincott Williams & Wilkins | 10.1213/ane.0b013e318165e208 | WOS:000254260200034 |
| Effect of heat- and steam-generating sheet on daily activities of living in patients with osteoarthritis of the knee: randomized prospective study | Journal of Orthopaedic Science: Official Journal of the Japanese Orthopaedic Association | Springer | 10.1007/s00776-008-1214-x | WOS:000256476800004 |
| A randomized clinical trial of the effects of isosorbide mononitrate on bone formation and resorption in post-menopausal women: a pilot study | Human Reproduction | Oxford University Press | 10.1093/humrep/dei487 | WOS:000236818600036 |
| High-dose chemotherapy with hematopoietic rescue as primary treatment for metastatic breast cancer: a randomized trial | Journal of Clinical Oncology : Official Journal of the American Society of Clinical Oncology | American Society of Clinical Oncology | 10.1200/JCO.1995.13.10.2483 | WOS:A1995RZ74400003 |
| Interferon alfa-2b, colchicine, and benzathine penicillin versus colchicine and benzathine penicillin in BehÃ§et's disease: a randomised trial | Lancet | Elsevier | 10.1016/S0140-6736(99)05131-4 | WOS:000085615400011 |

Table S3: Reasons of retraction of randomised controlled trials (with duplication)

|  | n |
| --- | --- |
| Investigation by Journal/Publisher | 31 |
| Investigation by Company/Institution | 26 |
| Concerns/Issues About Data | 24 |
| Falsification/Fabrication of Data | 24 |
| Misconduct by Author | 23 |
| Investigation by Third Party | 20 |
| Misconduct - Official Investigation/Finding | 19 |
| Plagiarism of Article | 15 |
| Lack of IRB/IACUC Approval | 12 |
| Error in Data | 11 |
| Objections by Third Party | 11 |
| Unreliable Data | 9 |
| Concerns/Issues About Results | 8 |
| Error in Methods | 8 |
| Ethical Violations by Author | 8 |
| Error in Analyses | 7 |
| Unreliable Results | 7 |
| Duplication of Article | 6 |
| Duplication of Data | 6 |
| Error in Results and/or Conclusions | 6 |
| Informed/Patient Consent - None/Withdrawn | 6 |
| Objections by Author(s) | 6 |
| False/Forged Authorship | 4 |
| Lack of Approval from Company/Institution | 4 |
| Original Data not Provided | 4 |
| Concerns/Issues About Authorship | 3 |
| Notice - Limited or No Information | 3 |
| Results Not Reproducible | 3 |
| Upgrade/Update of Prior Notice | 3 |
| Breach of Policy by Author | 2 |
| Euphemisms for Plagiarism | 2 |
| Plagiarism of Data | 2 |
| Plagiarism of Text | 2 |
| Author Unresponsive | 1 |
| Bias Issues or Lack of Balance | 1 |
| Complaints about Author | 1 |
| Concerns/Issues about Third Party Involvement | 1 |
| Conflict of Interest | 1 |
| Criminal Proceedings | 1 |
| Duplication of Image | 1 |
| Error by Third Party | 1 |
| Error in Image | 1 |
| Error in Text | 1 |
| Euphemisms for Misconduct | 1 |
| Lack of Approval from Author | 1 |
| Lack of Approval from Third Party | 1 |
| Miscommunication by Author | 1 |
| Miscommunication by Third Party | 1 |
| Publishing Ban | 1 |
| Retract and Replace | 1 |
| Salami Slicing | 1 |

Table S4. SR and CPG citing retracted RCT

| **title** | **url** | **SR_CPG** | **WOSID** |
| --- | --- | --- | --- |
| Medial subvastus versus the medial parapatellar approach for total knee replacement: a systematic review and meta-analysis of randomized controlled trials | 10.1302/2058-5241.3.170030 | SR | WOS:000428739000002 |
| Patellar non-eversion in primary TKA reduces the complication rate | 10.1007/s00167-015-3528-5 | SR | WOS:000371300400041 |
| Effect of Palliative Care on Quality of Life and Survival after Cardiopulmonary Resuscitation: A Systematic Review | 10.4103/ijpvm.IJPVM_191_18 | SR | WOS:000564355800003 |
| The Roles of ACE Inhibitors in Lower Extremity Peripheral Artery Disease | 10.1097/MJT.0000000000000011 | SR | WOS:000368122200003 |
| Supervised vs unsupervised exercise for intermittent claudication: A systematic review and meta-analysis | 10.1016/j.ahj.2015.03.009 | SR | WOS:000355213300023 |
| Society for Vascular Surgery practice guidelines for atherosclerotic occlusive disease of the lower extremities: Management of asymptomatic disease and claudication | 10.1016/j.jvs.2014.12.009 | CPG | WOS:000350219200002 |
| Consensus Document on Intermittent Claudication from the Central European Vascular Forum (CEVF)-3rd revision (2013) with the sharing of the Mediterranean League of Angiology and Vascular Surgery, and the North Africa and Middle East Chapter of International Union of Angiology | | CPG | WOS:000343531600005 |
| Angiotensin-Converting Enzyme Inhibitors for Intermittent Claudication Associated With Peripheral Arterial Disease | 10.1177/1060028013501803 | SR | WOS:000328147400021 |
| Treatment of hypertension in peripheral arterial disease | 10.1002/14651858.CD003075.pub3 | SR | WOS:000270687300033 |
| The effects of organic nitrates on osteoporosis: a systematic review | 10.1007/s00198-012-2262-9 | SR | WOS:000314889100002 |
| Nitroglycerin Ointment for the Prevention of Postmenopausal Osteoporosis | 10.1345/aph.1Q410 | SR | WOS:000298519200012 |
| Efficacy and safety of bone substitutes in lumbar spinal fusion: a systematic review and network meta-analysis of randomized controlled trials | 10.1007/s00586-019-06257-x | SR | WOS:000504137600002 |
| Minimally Invasive Versus Open Laminectomy/Discectomy, Transforaminal Lumbar, and Posterior Lumbar Interbody Fusions: A Systematic Review | 10.7759/cureus.1488 | SR | WOS:000453623000073 |
| Exploratory meta-analysis on dose-related efficacy and morbidity of bone morphogenetic protein in spinal arthrodesis surgery | 10.3171/2015.4.SPINE141086 | SR | WOS:000370915500019 |
| Exploring trajectories in dietary adequacy of the B vitamins folate, riboflavin, vitamins B-6 and B-12, with advancing older age: a systematic review | 10.1017/S0007114520004249 | SR | WOS:000671929700013 |
| The association of homocysteine, folate, vitamin B12, and vitamin B6 with fracture incidence in older adults: a systematic review and meta-analysis | 10.21037/atm-21-2514 | SR | WOS:000677659000001 |
| An investigation into the impact and implications of published papers from retracted research: systematic search of affected literature | 10.1136/bmjopen-2019-031909 | SR | WOS:000512882200249 |
| Interventions for preventing falls in older people in care facilities and hospitals | 10.1002/14651858.CD005465.pub4 | SR | WOS:000312255900036 |
| Effect of B Vitamin (Folate, B6, and B12) Supplementation on Osteoporotic Fracture and Bone Turnover Markers: A Meta-Analysis | | SR | WOS:000352364500002 |
| Guidelines for the Prevention of Stroke in Patients With Stroke and Transient Ischemic Attack A Guideline for Healthcare Professionals From the American Heart Association/American Stroke Association | 10.1161/STR.0000000000000024 | CPG | WOS:000285636400046 |
| ASPEN Clinical Guidelines: Nutrition Support of Neonatal Patients at Risk for Metabolic Bone Disease | 10.1177/0148607113487216 | CPG | WOS:000323896700005 |
| Broad-spectrum micronutrient formulas for the treatment of psychiatric symptoms: a systematic review | 10.1586/ERN.12.143 | SR | WOS:000314438400012 |
| Vitamin B12, Folate, Homocysteine, and Bone Health in Adults and Elderly People: A Systematic Review with Meta-Analyses | 10.1155/2013/486186 | SR | WOS:000214821100010 |
| Homocysteine level and risk of fracture: A meta-analysis and systematic review | 10.1016/j.bone.2012.05.024 | SR | WOS:000307617400009 |
| Systematic review of safety and tolerability of a complex micronutrient formula used in mental health | 10.1186/1471-244X-11-62 | SR | WOS:000290619500001 |
| A review of select vitamins and minerals used by postmenopausal women | 10.1016/j.maturitas.2010.06.003 | SR | WOS:000280944500007 |
| Interventions for preventing falls in older people in nursing care facilities and hospitals | 10.1002/14651858.CD005465.pub2 | SR | WOS:000274768700009 |
| Randomized clinical stroke rehabilitation trials in 2005 | 10.1007/s11064-006-9211-y | SR | WOS:000245134500027 |
| The role of hyperhomocysteinemia as well as folate, vitamin B-6 and B-12 deficiencies in osteoporosis a systematic review | 10.1515/CCLM.2007.362 | SR | WOS:000251964000008 |
| Vitamin D supplementation for prevention of mortality in adults | 10.1002/14651858.CD007470.pub3 | SR | WOS:000330508100031 |
| RETRACTED: Efficacy of Antiresorptive Agents for Preventing Fractures in Japanese Patients with an Increased Fracture Risk: Review of the Literature (Retracted article. See vol.34,pg.415,2017) | 10.2165/11597480-000000000-00000 | SR | WOS:000301829500003 |
| Interventions for preventing falls in older people living in the community | 10.1002/14651858.CD007146.pub3 | SR | WOS:000308828600027 |
| Vitamin D and prevention of falls in the elderly: a systematic review | | SR | WOS:000217104200011 |
| Pharmacologic treatment of low bone density or osteoporosis to prevent fractures: A clinical practice guideline from the American College of Physicians | 10.7326/0003-4819-149-6-200809160-00007 | CPG | WOS:000259230200005 |
| Systematic review: Comparative effectiveness of treatments to prevent fractures in men and women with low bone density or osteoporosis | 10.7326/0003-4819-148-3-200802050-00198 | SR | WOS:000252849700004 |
| Guidelines on muscle relaxants and reversal in anaesthesia | 10.1016/j.accpm.2020.01.005 | CPG | WOS:000517828500026 |
| Vaginal preparation with antiseptic solution before cesarean section for preventing postoperative infections | 10.1002/14651858.CD007892.pub7 | SR | WOS:000347645800045 |
| Vaginal Cleansing Before Cesarean Delivery A Systematic Review and Meta-analysis | 10.1097/AOG.0000000000002167 | SR | WOS:000408150300014 |
| Regular (ICSI) versus ultra-high magnification (IMSI) sperm selection for assisted reproduction | 10.1002/14651858.CD010167.pub3 | SR | WOS:000517176800014 |
| Live birth and miscarriage rate following intracytoplasmic morphologically selected sperm injection vs intracytoplasmic sperm injection: An updated systematic review and meta-analysis | 10.1111/aogs.13703 | SR | WOS:000484270900001 |
| Analysis of the effectiveness of assisted reproduction techniques: a systematic review | 10.23938/ASSN.0254 | SR | WOS:000432987500012 |
| Strategies to improve fertilisation rates with assisted conception: a systematic review | 10.1080/14647273.2017.1324182 | SR | WOS:000452041100003 |
| Summary of evidence-based nutritional recommendations of the Clinical Practice Guideline for the management of patients with Parkinson's disease | 10.20960/nh.287 | CPG | WOS:000388654600035 |
| The effectiveness of stretching for infants with congenital muscular torticollis | 10.1080/10833196.2019.1570704 | SR | WOS:000469002900002 |
| Effects of intraoperative high versus low inspiratory oxygen fraction (FiO(2)) on patient's outcome: A systematic review of evidence from the last 20 years | 10.1016/j.accpm.2020.07.019 | SR | WOS:000598914100035 |
| Safety of 80% vs 30-35% fraction of inspired oxygen in patients undergoing surgery: a systematic review and meta-analysis | 10.1016/j.bja.2018.11.026 | SR | WOS:000458513600014 |
| Effectiveness of 80% vs 30-35% fraction of inspired oxygen in patients undergoing surgery: an updated systematic review and meta-analysis | 10.1016/j.bja.2018.11.024 | SR | WOS:000458513600015 |
| Oxygen therapies and their effects on wound healing | 10.1111/wrr.12561 | SR | WOS:000416631700183 |
| Intraoperative modifiable risk factors of colorectal anastomotic leakage: Why surgeons and anesthesiologists should act together | 10.1016/j.ijsu.2016.09.098 | SR | WOS:000396458700031 |
| The effects of high perioperative inspiratory oxygen fraction for adult surgical patients | 10.1002/14651858.CD008884.pub2 | SR | WOS:000357606400040 |
| Sexual dysfunction in male individuals with spinal cord iniury: What do we know so far? | 10.1016/j.jocn.2019.07.038 | SR | WOS:000486093600004 |
| Clinical studies on low intensity extracorporeal shockwave therapy for erectile dysfunction: a systematic review and meta-analysis of randomised controlled trials | 10.1038/s41443-019-0117-z | SR | WOS:000467805000004 |
| Ramosetron for the prevention of postoperative nausea and vomiting (PONV): a meta-analysis | 10.4097/kjae.2011.61.5.405 | SR | WOS:000420593500010 |
| Lidocaine for reducing propofol-induced pain on induction of anaesthesia in adults | 10.1002/14651858.CD007874.pub2 | SR | WOS:000373285000032 |
| Prevention of pain on injection of propofol: systematic review and meta-analysis | 10.1136/bmj.d1110 | SR | WOS:000288698100004 |
| Efficacy and safety of flurbiprofen axetil in the prevention of pain on propofol injection: A systematic review and meta-analysis | | SR | WOS:000337258200001 |
| Efficacy and safety of flurbiprofen axetil on preemptive analgesia for Chinese surgical patients: A meta-analysis | 10.5897/AJPP11.805 | SR | WOS:000307973600002 |
| The efficacy of dexamethasone reducing postoperative pain and emesis after total knee arthroplasty: A systematic review and meta-analysis | 10.1016/j.ijsu.2018.02.043 | SR | WOS:000430456700026 |
| Preoperative intravenous glucocorticoids can decrease acute pain and postoperative nausea and vomiting after total hip arthroplasty A PRISMA-compliant meta-analysis | 10.1097/MD.0000000000008804 | SR | WOS:000417645700074 |
| Perioperative systemic steroid for rapid recovery in total knee and hip arthroplasty: a systematic review and meta-analysis of randomized trials | 10.1186/s13018-017-0601-4 | SR | WOS:000405006700001 |
| Perioperative systemic glucocorticoids in total hip and knee arthroplasty: A systematic review of outcomes | 10.1016/j.jor.2017.03.012 | SR | WOS:000406003600016 |
| Can intravenous steroid administration reduce postoperative pain scores following total knee arthroplasty? A meta-analysis | 10.1097/MD.0000000000007134 | SR | WOS:000403594100023 |
| A systematic review and meta-analysis of intravenous glucocorticoids for acute pain following total hip arthroplasty | 10.1097/MD.0000000000006872 | SR | WOS:000401188600054 |
| Low-dose dexamethasone during arthroplasty: what do we know about the risks? | 10.1302/2058-5241.1.000039 | SR | WOS:000441867300003 |
| Perioperative glucocorticoids in hip and knee surgery - benefit vs. harm? A review of randomized clinical trials | 10.1111/aas.12115 | SR | WOS:000321297200002 |
| Is protein the forgotten ingredient: Effects of higher compared to lower protein diets on cardiometabolic risk factors. A systematic review and meta-analysis of randomised controlled trials | 10.1016/j.atherosclerosis.2021.05.011 | SR | WOS:000668002800004 |
| Searching for the Antioxidant, Anti-Inflammatory, and Neuroprotective Potential of Natural Food and Nutritional Supplements for Ocular Health in the Mediterranean Population | 10.3390/foods10061231 | SR | WOS:000666512400001 |
| Nut consumption and type 2 diabetes risk: a systematic review and meta-analysis of observational studies | 10.1093/ajcn/nqaa358 | SR | WOS:000637368700025 |
| Mediterranean Diet and Telomere Length: A Systematic Review and Meta-Analysis | 10.1093/advances/nmaa079 | SR | WOS:000593351300009 |
| Effect of walnut consumption on markers of blood glucose control: a systematic review and meta-analysis | 10.1017/S0007114520001415 | SR | WOS:000564305000001 |
| A systematic review of the association between dietary patterns and health-related quality of life | 10.1186/s12955-020-01581-z | SR | WOS:000583271600001 |
| Bone loss, low height, and low weight in different populations and district: a meta-analysis between vegans and non-vegans | 10.29219/fnr.v64.3315 | SR | WOS:000569764100001 |
| Pharmacotherapeutic options for prediabetes | 10.1080/14656566.2020.1817381 | SR | WOS:000566997100001 |
| Impact of Nutrition on Telomere Health: Systematic Review of Observational Cohort Studies and Randomized Clinical Trials | 10.1093/advances/nmz107 | SR | WOS:000589649100009 |
| Obtaining evidence base for the development of Feel4Diabetes intervention to prevent type 2 diabetes - a narrative literature review | 10.1186/s12902-019-0468-y | SR | WOS:000521062000003 |
| Sociodemographic and lifestyle-related risk factors for identifying vulnerable groups for type 2 diabetes: a narrative review with emphasis on data from Europe | 10.1186/s12902-019-0463-3 | SR | WOS:000521062000002 |
| Reduction in saturated fat intake for cardiovascular disease | 10.1002/14551858.CD011737.pub3 | SR | WOS:000537031300050 |
| Improving type 2 diabetes mellitus glycaemic control through lifestyle modification implementing diet intervention: a systematic review and meta-analysis | 10.1007/s00394-019-02147-6 | SR | WOS:000499247000001 |
| Prevention of Type 2 Diabetes by Lifestyle Changes: A Systematic Review and Meta-Analysis | 10.3390/nu11112611 | SR | WOS:000502274600062 |
| Dietary fat intake and metabolic syndrome in adults: A systematic review | 10.1016/j.numecd.2019.05.055 | SR | WOS:000478962200001 |
| Role of diet in type 2 diabetes incidence: umbrella review of meta-analyses of prospective observational studies | 10.1136/bmj.l2368 | SR | WOS:000474836200001 |
| Mediterranean Diet and Cardiodiabesity: A Systematic Review through Evidence-Based Answers to Key Clinical Questions | 10.3390/nu11030655 | SR | WOS:000464361100003 |
| Prevention or Delay of Type 2 Diabetes: Standards of Medical Care in Diabetes-2019 | 10.2337/dc19-S003 | SR | WOS:000454291700006 |
| Olive Oil and Body Weight. Systematic Review and Meta-Analysis of Randomized Controlled Trials | | SR | WOS:000456322700001 |
| Glucose-Lowering Therapies for Cardiovascular Risk Reduction in Type 2 Diabetes Mellitus: State-of-the-Art Review | 10.1016/j.mayocp.2018.07.018 | SR | WOS:000448967300014 |
| The effects of the Mediterranean diet on rheumatoid arthritis prevention and treatment: a systematic review of human prospective studies | 10.1007/s00296-017-3912-1 | SR | WOS:000430547600004 |
| A Systematic Review of Behavioural Interventions Promoting Healthy Eating among Older People | 10.3390/nu10020128 | SR | WOS:000427540000018 |
| Diet and Men's Sexual Health | 10.1016/j.sxmr.2017.07.004 | SR | WOS:000496747100008 |
| Polyunsaturated fatty acids for the primary and secondary prevention of cardiovascular disease | 10.1002/14651858.CD012345.pub2 | SR | WOS:000455159600009 |
| The Effectiveness and Cost of Lifestyle Interventions Including Nutrition Education for Diabetes Prevention: A Systematic Review and Meta-Analysis | 10.1016/j.jand.2016.11.016 | SR | WOS:000395495400009 |
| AMERICAN ASSOCIATION OF CLINICAL ENDOCRINOLOGISTS AND AMERICAN COLLEGE OF ENDOCRINOLOGY COMPREHENSIVE CLINICAL PRACTICE GUIDELINES FOR MEDICAL CARE OF PATIENTS WITH OBESITY | 10.4158/EP161365.GL | SR | WOS:000384279500001 |
| The Effects of Breakfast Consumption and Composition on Metabolic Wellness with a Focus on Carbohydrate Metabolism | 10.3945/an.115.010314 | SR | WOS:000376163900019 |
| The Role of Energy, Nutrients, Foods, and Dietary Patterns in the Development of Gestational Diabetes Mellitus: A Systematic Review of Observational Studies | 10.2337/dc15-0540 | SR | WOS:000367331800018 |
| Nut consumption and risk of cancer and type 2 diabetes: a systematic review and meta-analysis | 10.1093/nutrit/nuv006 | SR | WOS:000359651100001 |
| Does cooking with vegetable oils increase the risk of chronic diseases?: a systematic review | 10.1017/S0007114514002931 | SR | WOS:000372517300006 |
| Effect of Mediterranean diet in diabetes control and cardiovascular risk modification: a systematic review | 10.3389/fpubh.2015.00069 | SR | WOS:000498912900068 |
| Role of Mediterranean diet in prevention and management of type 2 diabetes | 10.3760/cma.j.issn.0366-6999.20141358 | SR | WOS:000344724100023 |
| Mediterranean Diet and Cardiodiabesity: A Review | 10.3390/nu6093474 | SR | WOS:000342903700008 |
| Which diet for prevention of type 2 diabetes? A meta-analysis of prospective studies | 10.1007/s12020-014-0264-4 | SR | WOS:000344087600014 |
| Prevention of type 2 diabetes; a systematic review and meta-analysis of different intervention strategies | 10.1111/dom.12270 | SR | WOS:000340612400006 |
| Nut consumption and risk of type 2 diabetes, cardiovascular disease, and all-cause mortality: a systematic review and meta-analysis | 10.3945/ajcn.113.076109 | SR | WOS:000337862000029 |
| Nut consumption in relation to cardiovascular disease risk and type 2 diabetes: a systematic review and meta-analysis of prospective studies | 10.3945/ajcn.113.079152 | SR | WOS:000491255400002 |
| The effect of Mediterranean diet on the development of type 2 diabetes mellitus: A meta-analysis of 10 prospective studies and 136,846 participants | 10.1016/j.metabol.2014.04.010 | SR | WOS:000337715000004 |
| A Mediterranean diet improves HbA1c but not fasting blood glucose compared to alternative dietary strategies: a network meta-analysis | 10.1111/jhn.12138 | SR | WOS:000336216900011 |
| Effectiveness of dietary interventions among adults of retirement age: a systematic review and meta-analysis of randomized controlled trials | 10.1186/1741-7015-12-60 | SR | WOS:000335254500001 |
| Effects of Monounsaturated Fatty Acids on Glycaemic Control in Patients with Abnormal Glucose Metabolism: A Systematic Review and Meta-Analysis | 10.1159/000331214 | SR | WOS:000296422500005 |
| Mesenchymal Stem Cells in Cardiac Repair: Effects on Myocytes, Vasculature, and Fibroblasts | 10.1016/j.clinthera.2020.08.010 | SR | WOS:000596677000005 |
| Utilisation of real-world data from heart failure registries in OECD countries - A systematic review | 10.1016/j.ijcha.2018.02.006 | SR | WOS:000436325100018 |
| Cellular therapies for chronic ischemic heart failure | 10.1016/j.hjc.2018.01.010 | SR | WOS:000440049300003 |
| Adult Stem Cell Therapy for Cardiac Repair in Patients After Acute Myocardial Infarction Leading to Ischemic Heart Failure: An Overview of Evidence from the Recent Clinical Trials | 10.2174/1573403X13666170502103833 | SR | WOS:000415722900007 |
| Cardiac Stem Cell Treatment in Myocardial Infarction: A Systematic Review and Meta-Analysis of Preclinical Studies | 10.1161/CIRCRESAHA.115.307676 | SR | WOS:000374779300007 |
| Applications of regenerative medicine in organ transplantation | 10.4103/0975-7406.160013 | SR | WOS:000219305300004 |
| Similar Effect of Autologous and Allogeneic Cell Therapy for Ischemic Heart Disease Systematic Review and Meta-Analysis of Large Animal Studies | 10.1161/CIRCRESAHA.116.304872 | SR | WOS:000347052800015 |
| 2013 ACCF/AHA Guideline for the Management of ST-Elevation Myocardial Infarction A Report of the American College of Cardiology Foundation/American Heart Association Task Force on Practice Guidelines | 10.1016/j.jacc.2012.11.019 | SR | WOS:000313835300002 |
| Adult Bone Marrow Cell Therapy Improves Survival and Induces Long-Term Improvement in Cardiac Parameters A Systematic Review and Meta-Analysis | 10.1161/CIRCULATIONAHA.111.086074 | SR | WOS:000306977500013 |
| Effects of dexmedetomidine on perioperative stress, inflammation, and immune function: systematic review and meta-analysis | 10.1016/j.bja.2019.07.027 | SR | WOS:000496915500011 |
| Denosumab for Effective Tumor Size Reduction in Patients With Giant Cell Tumors of the Bone: A Systematic Review and Meta-Analysis | 10.1177/1073274820934822 | SR | WOS:000570936200001 |
| The Relationship of Tree Nuts and Peanuts with Adiposity Parameters: A Systematic Review and Network Meta-Analysis | 10.3390/nu13072251 | SR | WOS:000676535800001 |
| Behavioral Counseling to Promote a Healthy Diet and Physical Activity for Cardiovascular Disease Prevention in Adults With Cardiovascular Risk Factors Updated Evidence Report and Systematic Review for the US Preventive Services Task Force | 10.1001/jama.2020.17108 | SR | WOS:000596035700024 |
| The Effects of Diets Enriched in Monounsaturated Oleic Acid on the Management and Prevention of Obesity: A Systematic Review of Human Intervention Studies | 10.1093/advances/nmaa013 | SR | WOS:000593344900008 |
| Inverse association of long-term nut consumption with weight gain and risk of overweight/obesity: a systematic review | 10.1016/j.nutres.2019.04.001 | SR | WOS:000496609400001 |
| Central obesity and the Mediterranean diet: A systematic review of intervention trials | 10.1080/10408398.2017.1351917 | SR | WOS:000458800200002 |
| Progestogens for preventing miscarriage: a network meta-analysis | 10.1002/14651858.CD013792.pub2 | SR | WOS:000646037100009 |
| Treatment efficacy for idiopathic recurrent pregnancy loss - a systematic review and meta-analyses | 10.1111/aogs.13352 | SR | WOS:000439485900003 |
| Recurrent Miscarriage: Diagnostic and Therapeutic Procedures. Guideline of the DGGG, OEGGG and SGGG (S2k-Level, AWMF Registry Number 015/050) | 10.1055/a-0586-4568 | CPG | WOS:000432255000010 |
| Care prior to and during subsequent pregnancies following stillbirth for improving outcomes | 10.1002/14651858.CD012203.pub2 | SR | WOS:000455302700006 |
| Efficacy and safety of ramosetron versus ondansetron for postoperative nausea and vomiting after general anesthesia: a meta-analysis of randomized clinical trials | 10.2147/DDDT.S80407 | SR | WOS:000353354900001 |
| Effect of intra-articular alpha-agonists on post-operative outcomes following arthroscopic knee surgery: A systematic review and meta-analysis | 10.1016/j.egja.2017.02.004 | SR | WOS:000414326400011 |
| RETRACTED: Efficacy of dexmedetomidine on postoperative nausea and vomiting: a meta-analysis of randomized controlled trials(Retracted article. See vol.9, pg.20465,2016) | | SR | WOS:000365271900025 |
| Dexamethasone Is Superior to Dexmedetomidine as a Perineural Adjunct for Supraclavicular Brachial Plexus Block: Systematic Review and Indirect Meta-analysis | 10.1213/ANE.0000000000003860 | SR | WOS:000460108900028 |
| Efficacy and safety of dexmedetomidine in peripheral nerve blocks: A meta-analysis and trial sequential analysis | 10.1097/EJA.0000000000000870 | SR | WOS:000456268000005 |
| Investigating the Efficacy of Dexmedetomidine as an Adjuvant to Local Anesthesia in Brachial Plexus Block A Systematic Review and Meta-Analysis of 18 Randomized Controlled Trials | 10.1097/AAP.0000000000000564 | SR | WOS:000394514600003 |
| Evidence basis for using perineural dexmedetomidine to enhance the quality of brachial plexus nerve blocks: a systematic review and meta-analysis of randomized controlled trials | 10.1093/bja/aew411 | SR | WOS:000395324500006 |
| Effects of dexmedetomidine as a local anesthetic adjuvant for brachial plexus block: a systematic review and meta-analysis | | SR | WOS:000396503400034 |
| The Efficacy of Hypotensive Agents on Intraoperative Bleeding and Recovery Following General Anesthesia for Nasal Surgery: A Network Meta-Analysis | 10.21053/ceo.2020.00584 | SR | WOS:000646218200009 |
| Use of alpha(2)-Adrenergic Agonists to Improve Surgical Field Visibility in Endoscopy Sinus Surgery: A Systematic Review of Randomised Controlled Trials | 10.1016/j.clinthera.2017.11.010 | SR | WOS:000423137800015 |
| Comparative Study of the Adverse Events Associated With Adjuvant Use of Dexmedetomidine and Clonidine in Local Anesthesia | 10.3389/fmed.2021.602966 | SR | WOS:000670573100001 |
| Post-operative pain management modalities employed in clinical trials for adult patients in LMIC; a systematic review | 10.1186/s12871-021-01375-w | SR | WOS:000654174900001 |
| Research Advances on Health Effects of Edible Artemisia Species and Some Sesquiterpene Lactones Constituents | 10.3390/foods10010065 | SR | WOS:000610211400001 |
| Artemisia afra, a controversial herbal remedy or a treasure trove of new drugs? | 10.1016/j.jep.2019.112127 | SR | WOS:000486357500003 |
| The comparative effectiveness of 55 interventions in obese patients with polycystic ovary syndrome: A network meta-analysis of 101 randomized trials | 10.1371/journal.pone.0254412 | SR | WOS:000678122000040 |
| The function of metformin in endometrial receptivity (ER) of patients with polycyclic ovary syndrome (PCOS): a systematic review and meta-analysis | 10.1186/s12958-021-00772-7 | SR | WOS:000661490900001 |
| Metformin for ovulation induction (excluding gonadotrophins) in women with polycystic ovary syndrome | 10.1002/14651858.CD013505 | SR | WOS:000505243400033 |
| Evidence summaries and recommendations from the international evidence-based guideline for the assessment and management of polycystic ovary syndrome: assessment and treatment of infertility | 10.1093/hropen/hoy021 | SR | WOS:000661872500001 |
| Aromatase inhibitors (letrozole) for subfertile women with polycystic ovary syndrome | 10.1002/14651858.CD010287.pub3 | SR | WOS:000348966700009 |
| Role of metformin for ovulation induction in infertile patients with polycystic ovary syndrome (PCOS): a guideline | 10.1016/j.fertnstert.2017.06.026 | SR | WOS:000408749600012 |
| Combined metformin-clomiphene in clomiphene-resistant polycystic ovary syndrome: a systematic review and meta-analysis of randomized controlled trials | 10.1111/aogs.12673 | SR | WOS:000359086600003 |
| Polycystic ovary syndrome: chemical pharmacotherapy | 10.1517/14656566.2015.1047344 | SR | WOS:000355893300009 |
| gg Aromatase inhibitors for subfertile women with polycystic ovary syndrome | 10.1002/14651858.CD010287.pub2 | SR | WOS:000332082900038 |
| Aromatase inhibitors for PCOS: a systematic review and meta-analysis | 10.1093/humupd/dms003 | SR | WOS:000303162500006 |
| Assessment and management of polycystic ovary syndrome: summary of an evidence-based guideline | 10.5694/mja11.10915 | SR | WOS:000295543300001 |
| Induction of ovulation | 10.1016/S0368-2315(10)70032-6 | SR | WOS:000285699800008 |
| Higher ovulation rate with letrozole as compared with clomiphene citrate in infertile women with polycystic ovary syndrome: a systematic review and meta-analysis | 10.1007/s42000-021-00289-z | SR | WOS:000654131500001 |
| Endometrial injury for pregnancy following sexual intercourse or intrauterine insemination | 10.1002/14651858.CD011424.pub3 | SR | WOS:000381106800038 |
| Letrozole versus laparoscopic ovarian drilling in clomiphene citrate-resistant women with polycystic ovary syndrome: a systematic review and meta-analysis of randomized controlled trials | 10.1186/s12958-019-0461-3 | SR | WOS:000458542600001 |
| Systematic review and meta-analysis of letrozole and clomiphene citrate in polycystic ovary syndrome | 10.1016/j.mefs.2018.03.008 | SR | WOS:000443739200001 |
| Effect of clomiphene citrate on endometrial thickness, ovulation, pregnancy and live birth in anovulatory women: systematic review and meta-analysis | 10.1002/uog.18933 | SR | WOS:000428231900009 |
| Treatment strategies for women with WHO group II anovulation: systematic review and network meta-analysis | 10.1136/bmj.j138 | SR | WOS:000393449700002 |
| Letrozole versus clomiphene citrate in polycystic ovary syndrome: systematic review and meta-analysis | 10.3109/09513590.2015.1096337 | SR | WOS:000366349300001 |
| Efficacy of luteal phase support with vaginal progesterone in intrauterine insemination: a systematic review and meta-analysis | 10.1007/s10815-013-0127-6 | SR | WOS:000330987600012 |
| Meta-analysis of letrozole versus clomiphene citrate in polycystic ovary syndrome | 10.1016/j.rbmo.2011.03.024 | SR | WOS:000303044300011 |
| Use of letrozole in assisted reproduction: a systematic review and meta-analysis | 10.1093/humupd/dmn033 | SR | WOS:000260152100005 |
| Comparison of clomiphene and letrozole for superovulation in patients with unexplained infertility undergoing intrauterine insemination A systematic review and meta-analysis | 10.1097/MD.0000000000021006 | SR | WOS:000562693800015 |
| Evidence-based treatments for couples with unexplained infertility: a guideline | 10.1016/j.fertnstert.2019.10.014 | CPG | WOS:000517758400009 |
| The management of unexplained infertility: an evidence-based guideline from the Canadian Fertility and Andrology Society | 10.1016/j.rbmo.2019.05.023 | CPG | WOS:000488774100012 |
| Letrozole Compared With Clomiphene Citrate for Unexplained Infertility A Systematic Review and Meta-analysis | 10.1097/AOG.0000000000003105 | SR | WOS:000334672400006 |
| No. 362-Ovulation Induction in Polycystic Ovary Syndrome | 10.1016/j.jogc.2017.12.004 | CPG | WOS:000436555100019 |
| Endometrial thickness in women undergoing IUI with ovarian stimulation. How thick is too thin? A systematic review and meta-analysis | 10.1093/humrep/dex035 | SR | WOS:000401012300006 |
| Letrozole versus clomiphene citrate for unexplained infertility: A systematic review and meta-analysis | 10.1111/jog.12393 | SR | WOS:000334672400006 |
| Ovulation Induction in Polycystic Ovary Syndrome No. 242, May 2010 | 10.1016/j.ijgo.2010.07.001 | CPG | WOS:000282735100027 |
| Ovulation Induction in Polycystic Ovary Syndrome | 10.1016/S1701-2163(16)34504-2 | CPG | WOS:000443260400012 |
| Chinese Herbal Medicine and Clomiphene Citrate for Anovulation: A Meta-Analysis of Randomized Controlled Trials | 10.1089/acm.2010.0254 | SR | WOS:000290784100004 |
| Aromatase Inhibitors for Ovulation and Pregnancy in Polycystic Ovary Syndrome | 10.1345/aph.1M096 | SR | WOS:000268512700021 |
| Is There a Role for Bismuth in Diarrhea Management? | 10.5041/RMMJ.10422 | SR | WOS:000609802100002 |
| Roux-en-Y gastric bypass versus one anastomosis-mini gastric bypass as a rescue procedure following failed restrictive bariatric surgery. A systematic review of literature with metanalysis | 10.1007/s13304-020-00938-9 | SR | WOS:000619762900001 |
| Areas of Non-Consensus Around One Anastomosis/Mini Gastric Bypass (OAGB/MGB): A Narrative Review | 10.1007/s11695-021-05276-2 | SR | WOS:000618908500002 |
| Laparoscopic Sleeve Gastrectomy Versus Laparoscopic Roux-en-Y Gastric Bypass A Systematic Review and Meta-analysis of Weight Loss, Comorbidities, and Biochemical Outcomes From Randomized Controlled Trials | 10.1097/SLA.0000000000003671 | SR | WOS:000613348700026 |
| Five-Year Weight Loss Outcomes in Laparoscopic Vertical Sleeve Gastrectomy (LVSG) Versus Laparoscopic Roux-en-Y Gastric Bypass (LRYGB) Procedures: A Systematic Review and Meta-Analysis of Randomized Controlled Trials | 10.1097/SLE.0000000000000834 | SR | WOS:000599708300015 |
| Bariatric Surgery and Type 2 Diabetes Mellitus: Assessing Factors Leading to Remission. A Systematic Review | 10.7759/cureus.9973 | SR | WOS:000561740100024 |
| Comparison of the effect of Roux-en-Y gastric bypass and sleeve gastrectomy on remission of type 2 diabetes: A systematic review and meta-analysis of randomized controlled trials | 10.1111/obr.13011 | SR | WOS:000533423200001 |
| Clinical practice guidelines of the European Association for Endoscopic Surgery (EAES) on bariatric surgery: update 2020 endorsed by IFSO-EC, EASO and ESPCOP | 10.1007/s00464-020-07555-y | SR | WOS:000528325600005 |
| One Anastomosis Gastric Bypass Versus Roux-en-Y Gastric Bypass for Obesity: a Systematic Review and Meta-Analysis of Randomized Clinical Trials | 10.1007/s11695-019-04288-3 | SR | WOS:000520712600005 |
| In Terms of Nutrition, the Most Suitable Method for Bariatric Surgery: Laparoscopic Sleeve Gastrectomy or Roux-en-Y Gastric Bypass? A Systematic Review and Meta-analysis | 10.1007/s11695-020-04488-2 | SR | WOS:000516354900003 |
| Laparoscopic Roux-en-Y gastric bypass versus laparoscopic sleeve gastrectomy for 5-year hypertension remission in obese patients: a systematic review and meta-analysis | 10.1097/HJH.0000000000002255 | SR | WOS:000524989300001 |
| Clinical Outcomes of One Anastomosis Gastric Bypass Versus Sleeve Gastrectomy for Morbid Obesity | 10.1007/s11695-019-04303-7 | SR | WOS:000500460200003 |
| Procedure and patient selection in bariatric and metabolic surgery | 10.23736/S0026-4733.19.08121-5 | SR | WOS:000500788100007 |
| One Anastomosis Gastric Bypass Versus Roux-en-Y Gastric Bypass for Morbid Obesity: an Updated Meta-Analysis | 10.1007/s11695-019-04005-0 | SR | WOS:000483706700002 |
| Mid-long-term Revisional Surgery After Sleeve Gastrectomy: a Systematic Review and Meta-analysis | 10.1007/s11695-019-03842-3 | SR | WOS:000468846000040 |
| Mesenchymal Stromal Cell-Conditioned Medium for Skin Diseases: A Systematic Review | 10.3389/fcell.2021.654210 | SR | WOS:000681621000001 |
| Fractional radiofrequency in the treatment of skin aging: an evidence-based treatment protocol | 10.1080/14764172.2019.1674448 | SR | WOS:000501970800001 |
| Microneedling in All Skin Types: A Review | | SR | WOS:000398431600002 |
| Microneedling: A Comprehensive Review | 10.1097/DSS.0000000000000924 | SR | WOS:000398047200001 |
| The Effect of Oral Magnesium Supplementation on Inflammatory Biomarkers in Adults: A Comprehensive Systematic Review and Dose-response Meta-analysis of Randomized Clinical Trials | 10.1007/s12011-021-02783-2 | SR | WOS:000663285000001 |
| The effects of vitamin and mineral supplementation on women with gestational diabetes mellitus | 10.1186/s12902-021-00712-x | SR | WOS:000653763200001 |
| Crosstalk of Magnesium and Serum Lipids in Dyslipidemia and Associated Disorders: A Systematic Review | 10.3390/nu13051411 | SR | WOS:000662341300001 |
| Effects of Nutritional Strategies on Glucose Homeostasis in Gestational Diabetes Mellitus: A Systematic Review and Network Meta-Analysis | 10.1155/2020/6062478 | SR | WOS:000522223100001 |
| Serum Magnesium Levels in Preterm Infants Are Higher Than Adult Levels: A Systematic Literature Review and Meta-Analysis | 10.3390/nu9101125 | SR | WOS:000414629900081 |
| Effect of magnesium supplementation on insulin resistance in humans: A systematic review | 10.1016/j.nut.2017.01.009 | SR | WOS:000402224000011 |
| Effect of Magnesium Supplementation on Plasma C-reactive Protein Concentrations: A Systematic Review and Meta-Analysis of Randomized Controlled Trials | 10.2174/1381612823666170525153605 | SR | WOS:000416930000012 |
| Effect of magnesium supplementation on glucose metabolism in people with or at risk of diabetes: a systematic review and meta-analysis of double-blind randomized controlled trials | 10.1038/ejcn.2016.154 | SR | WOS:000391349700003 |
| Changes in anthropometric and blood 25-hydroxyvitamin D measurements in antenatal vitamin supplemented gestational diabetes mellitus patients: a systematic review and meta-analysis of randomized controlled trials | 10.4274/jtgga.galenos.2021.2020.0197 | SR | WOS:000692172500010 |
| The effects of vitamin D supplementation on glycemic control and maternal-neonatal outcomes in women with established gestational diabetes mellitus: A systematic review and meta-analysis | 10.1016/j.clnu.2020.12.016 | SR | WOS:000654716700031 |
| Dietary supplementation for gestational diabetes prevention and management: a meta-analysis of randomized controlled trials | 10.1007/s00404-021-06023-9 | SR | WOS:000630979600001 |
| The Molecular Mechanisms by Which Vitamin D Prevents Insulin Resistance and Associated Disorders | 10.3390/ijms21186644 | SR | WOS:000582029800001 |
| Maternal Vitamin D Levels During Pregnancy and Their Effects on Maternal-Fetal Outcomes: A Systematic Review | 10.1016/j.jogc.2019.09.013 | SR | WOS:000569829500011 |
| A comparison of the risk of cesarean section in gestational diabetes mellitus patients supplemented antenatally with vitamin D containing supplements versus placebo: A systematic review and meta-analysis of double-blinded randomized controlled trials | 10.4274/jtgga.galenos.2020.2019.0164 | SR | WOS:000566719700010 |
| The risk of morbidities in newborns of antenatal vitamin D supplemented gestational diabetes mellitus patients | | SR | WOS:000564641400002 |
| Vitamin D supplementation and incident preeclampsia: A systematic review and meta-analysis of randomized clinical trials | 10.1016/j.clnu.2019.08.015 | SR | WOS:000536918600012 |
| Vitamin D Supplementation during Pregnancy: An Evidence Analysis Center Systematic Review and Meta-Analysis | 10.1016/j.jand.2019.07.002 | SR | WOS:000531565300013 |
| A systematic review and meta-analysis of the effect of Vitamin D-fortified food on glycemic indices | 10.1002/biof.1632 | SR | WOS:000550047300001 |
| Nutritional Gaps and Supplementation in the First 1000 Days | 10.3390/nu11122891 | SR | WOS:000506917800053 |
| The Effect of Vitamin D Supplementation on Glycaemic Control in Women with Gestational Diabetes Mellitus: A Systematic Review and Meta-Analysis of Randomised Controlled Trials | 10.3390/ijerph16101716 | SR | WOS:000470967500050 |
| Efficacy of vitamin D supplementation in gestational diabetes mellitus: Systematic review and meta-analysis of randomized trials | 10.1371/journal.pone.0213006 | SR | WOS:000462000400006 |
| Vitamin D and diabetic foot ulcer: a systematic review and meta-analysis | 10.1038/s41387-019-0078-9 | SR | WOS:000562693800009 |
| The effects of vitamin D supplementation on indices of glycemic control in Iranian diabetics: A systematic review and meta-analysis | 10.1016/j.ctcp.2018.12.009 | SR | WOS:000457640000043 |
| The effect of vitamin D supplementation on oxidative stress parameters: A systematic review and meta-analysis of clinical trials | 10.1016/j.phrs.2018.11.011 | SR | WOS:000458709000013 |
| Vitamin D and gestational diabetes mellitus: a systematic review based on data free of Hawthorne effect | 10.1111/1471-0528.15060 | SR | WOS:000433566700004 |
| The Effects of Vitamin D Supplementation on Biomarkers of Inflammation and Oxidative Stress in Diabetic Patients: A Systematic Review and Meta-Analysis of Randomized Controlled Trials | 10.1055/a-0630-1303 | SR | WOS:000434670100001 |
| The Effects of Synbiotic Supplementation on GlucoseMetabolism and Lipid Profiles in Patients with Diabetes: a Systematic Review and Meta-Analysis of Randomized Controlled Trials | 10.1007/s12602-017-9299-1 | SR | WOS:000439456000023 |
| Effects of Vitamin D Deficiency on Incidence Risk of Gestational Diabetes Mellitus: A Systematic Review and Meta-analysis | 10.3389/fendo.2018.00007 | SR | WOS:000423803700001 |
| Maternal and Neonatal Metabolic Outcomes of Vitamin D Supplementation in Gestational Diabetes Mellitus: A Systematic Review and Meta-Analysis | 10.1159/000491643 | SR | WOS:000444745500008 |
| Calcium and Vitamin D Supplementation for Prevention of Preeclampsia: A Systematic Review and Network Meta-Analysis | 10.3390/nu9101141 | SR | WOS:000414629900097 |
| Impact of vitamin D supplementation on endothelial and inflammatory markers in adults: A systematic review | 10.1016/j.jsbmb.2017.01.015 | SR | WOS:000412249700043 |
| The Effects of Vitamin D Supplementation on Glucose Metabolism and Lipid Profiles in Patients with Gestational Diabetes: A Systematic Review and Meta-Analysis of Randomized Controlled Trials | 10.1055/s-0043-115225 | SR | WOS:000410529100001 |
| THE RELATIONSHIP BETWEEN VITAMIN D AND GESTATIONAL DIABETES-A REVIEW ARTICLE | 10.5281/zenodo.843525 | SR | WOS:000411335600012 |
| Non-skeletal health effects of vitamin D supplementation: A systematic review on findings from meta-analyses summarizing trial data | 10.1371/journal.pone.0180512 | SR | WOS:000405464100072 |
| Vitamin D and histologic severity of nonalcoholic fatty liver disease: A systematic review and meta-analysis | 10.1016/j.dld.2017.02.003 | SR | WOS:000406221000005 |
| Impact of Probiotic Administration on Serum C-Reactive Protein Concentrations: Systematic Review and Meta-Analysis of Randomized Control Trials | 10.3390/nu9010020 | SR | WOS:000396465500020 |
| Vitamin D supplementation during pregnancy: Updated meta-analysis on maternal outcomes | 10.1016/j.jsbmb.2016.02.008 | SR | WOS:000388157000026 |
| Vitamin D supplementation for women during pregnancy | 10.1002/14651858.CD008873.pub3 | SR | WOS:000374404200033 |
| Effect of Vitamin D Supplementation on Blood Pressure A Systematic Review and Meta-analysis Incorporating Individual Patient Data | 10.1001/jamainternmed.2015.0237 | SR | WOS:000356178400013 |
| Vitamin D and pregnancy outcomes | 10.1097/GCO.0000000000000117 | SR | WOS:000344982000003 |
| Mind-Body Interventions in Late-Life Mental Illnesses and Cognitive Disorders: A Narrative Review | 10.1016/j.jagp.2018.10.020 | SR | WOS:000462591700013 |
| Pharmacotherapy for dissociative disorders: A systematic review | 10.1016/j.psychres.2019.112529 | SR | WOS:000497252200010 |
| Lamotrigine Uses in Psychiatric Practice | 10.1097/MJT.0000000000000535 | SR | WOS:000467746200014 |
| Efficacy and safety of immunomodulatory drugs in patients with anterior uveitis: A systematic literature review | 10.1097/MD.0000000000008045 | SR | WOS:000415100000031 |
| The Treatment of Chronic Recurrent Oral Aphthous Ulcers | 10.3238/arztebl.2014.0665 | SR | WOS:000344902500001 |
| Development of consensus statements for the diagnosis and management of intestinal Behcet's disease using a modified Delphi approach | 10.1007/s00535-007-2090-4 | CPG | WOS:000249577300005 |
| The use of interferon alpha in Behcet disease: Review of the literature | 10.1053/S0049-0172(03)00167-7 | SR | WOS:000220932800005 |
| High dose chemotherapy and autologous bone marrow or stem cell transplantation versus conventional chemotherapy for women with metastatic breast cancer | 10.1002/14651858.CD003142.pub2 | SR | WOS:000232202500118 |
| A systematic overview of chemotherapy effects in breast cancer | 10.1080/02841860151116349 | SR | WOS:000169253000010 |
| Systematic reviews of chemotherapy and endocrine therapy in metastatic breast cancer | 10.1053/ctrv.1999.0161 | SR | WOS:000087350900001 |
| Autologous stem cell transplantation for malignancy: a systematic review of the literature | 10.1046/j.1365-2257.2000.00270.x | SR | WOS:000087420200001 |
| Chemotherapy-induced anemia in adults: Incidence and treatment | 10.1093/jnci/91.19.1616 | SR | WOS:000082932400009 |
| An overview of evidence-based management of hepatocellular carcinoma: A meta-analysis | 10.4103/0973-1482.92023 | SR | WOS:000299473200016 |
| Systematic review and meta-analysis of survival and disease recurrence after radiofrequency ablation for hepatocellular carcinoma | 10.1002/bjs.7669 | SR | WOS:000294355000006 |
| The Current Role of Radiofrequency Ablation in the Management of Hepatocellular Carcinoma A Systematic Review | 10.1097/SLA.0b013e31818eec29 | SR | WOS:000262219300005 |
| First-line tandem high-dose chemotherapy and autologous stem cell transplantation versus single high-dose chemotherapy and autologous stem cell transplantation in multiple myeloma, a systematic review of controlled studies | 10.1002/14651858.CD004626.pub3 | SR | WOS:000310016900004 |
| International Myeloma Working Group guidelines for the management of multiple myeloma patients ineligible for standard high-dose chemotherapy with autologous stem cell transplantation | 10.1038/leu.2009.122 | CPG | WOS:000270816300005 |
| Tandem Versus Single Autologous Hematopoietic Cell Transplantation for the Treatment of Multiple Myeloma: A Systematic Review and Meta-analysis | 10.1093/jnci/djn439 | SR | WOS:000313957300002 |
| A meta-analysis and systematic review of thalidomide for patients with previously untreated multiple myeloma | 10.1016/j.ctrv.2008.02.003 | SR | WOS:000258393900004 |
| Perioperative angiotensin-converting enzyme inhibitors or angiotensin II type 1 receptor blockers for preventing mortality and morbidity in adults | 10.1002/14651858.CD009210.pub2 | SR | WOS:000374404200037 |
| Cytotoxic and hormonal treatment for metastatic breast cancer: A systematic review of published randomized trials involving 31,510 women | 10.1200/JCO.1998.16.10.3439 | SR | WOS:000076347100032 |
| Much ado about not ... enough data: High-dose chemotherapy with autologous stem cell rescue for breast cancer | 10.1093/jnci/90.3.200 | SR | WOS:000072121500007 |
| Clinical impact of chemotherapy dose escalation in patients with hematologic malignancies and solid tumors | 10.1200/JCO.1997.15.8.2981 | SR | WOS:A1997XN95400027 |
| Consensus on the use of neutrophil-stimulating hematopoietic growth factors in clinical practice: An international viewpoint | 10.1016/S0924-8579(97)00022-8 | CPG | WOS:A1997XH82300008 |
| Nonantimuscarinic treatment for overactive bladder: a systematic review | 10.1016/j.ajog.2016.01.156 | SR | WOS:000378630000006 |
| Hematoma Risks of Nonsteroidal Anti-inflammatory Drugs Used in Plastic Surgery Procedures A Systematic Review and Meta-analysis | 10.1097/SAP.0000000000001898 | SR | WOS:000473359700015 |
| Perioperative Administration of Selective Cyclooxygenase-2 Inhibitors for Postoperative Pain Management in Patients After Total Knee Arthroplasty | 10.1016/j.arth.2012.04.008 | SR | WOS:000314440600002 |
| Pharmacotherapy for the prevention of chronic pain after surgery in adults | 10.1002/14651858.CD008307.pub2 | SR | WOS:000322568300023 |
| Paracetamol and selective and non-selective nonsteroidal anti-inflammatory drugs (NSAIDs) for the reduction of morphine-related side effects after major surgery:a systematic review | 10.3310/hta14170 | SR | WOS:000277278600001 |
| Dual Renin-Angiotensin-Aldosterone Blockade: Promises and Pitfalls | 10.1007/s11906-014-0511-3 | SR | WOS:000351563600001 |
| Angiotensin converting enzyme (ACE) inhibitors versus angiotensin receptor blockers for primary hypertension | 10.1002/14651858.CD009096.pub2 | SR | WOS:000209700700005 |
| Efficacy and safety of dual blockade of the renin-angiotensin system: meta-analysis of randomised trials | 10.1136/bmj.f360 | SR | WOS:000314509400001 |
| The Role of ARBs Alone or with HCTZ in the Treatment of Hypertension and Prevention of Cardiovascular and Renal Complications | 10.3810/pgm.2012.03.2535 | SR | WOS:000303420500004 |
| Angiotensin-converting enzyme inhibitors, angiotensin receptor blockers and combined therapy in patients with micro- and macroalbuminuria and other cardiovascular risk factors: a systematic review of randomized controlled trials | 10.1093/ndt/gfq792 | SR | WOS:000295231600018 |
| Early change in proteinuria as a surrogate outcome in kidney disease progression: a systematic review of previous analyses and creation of a patient-level pooled dataset | 10.1093/ndt/gfq525 | SR | WOS:000287746500013 |
| Angiotensin-converting enzyme inhibitors and angiotensin receptor blockers for adults with early (stage 1 to 3) non-diabetic chronic kidney disease | 10.1002/14651858.CD007751.pub2 | SR | WOS:000295674300045 |
| European guidelines on the management of hypertension: The European Society of Hypertension position statement (2009) | | CPG | WOS:000433849000001 |
| Present and Prospective Clinical Therapeutic Options for the Elderly Patient with Hypertension | | SR | WOS:000215767200060 |
| Renal protective effect of RAAS blockade across the renal continuum, with a review of the efficacy and safety of valsartan | 10.1185/03007990903328231 | SR | WOS:000273417800013 |
| Management of people with intermediate-stage hepatocellular carcinoma: an attempted network meta-analysis | 10.1002/14651858.CD011649.pub2 | SR | WOS:000400761200057 |
| Radiofrequency ablation plus chemoembolization versus radiofrequency ablation alone for hepatocellular carcinoma: A systematic review and meta-analysis | 10.1016/j.clinre.2015.07.008 | SR | WOS:000377426200012 |
| Comparative Efficacy of Interventional Therapies for Early-stage Hepatocellular Carcinoma  A PRISMA-compliant Systematic Review and Network Meta-analysis | 10.1097/MD.0000000000003185 | SR | WOS:000376924500021 |
| Comparative analysis of current guidelines for the treatment of hepatocellular carcinoma | 10.2217/hep-2015-0006 | SR | WOS:000398947300004 |
| EFSUMB Guidelines on Interventional Ultrasound (INVUS), Part III Abdominal Treatment Procedures (Long Version) | 10.1055/s-0035-1553917 | CPG | WOS:000370626700001 |
| Radiofrequency ablation with or without transarterial chemoembolization for hepatocellular carcinoma: A systematic review and meta-analysis | 10.13105/wjma.v3.i6.295 | SR | WOS:000367893600005 |
| Efficacy of percutaneous radiofrequency ablation for the treatment of hepatocellular carcinoma | 10.4238/2015.December.22.24 | SR | WOS:000371587400073 |
| Combination of radiofrequency ablation with transarterial chemoembolization for hepatocellular carcinoma: an up-to-date meta-analysis | 10.1007/s13277-014-1976-z | SR | WOS:000341883500019 |
| Comparative effectiveness of radiofrequency ablation with or without transarterial chemoembolization for hepatocellular carcinoma | 10.1007/s13277-013-1349-z | SR | WOS:000333536300115 |
| Meta-analysis of radiofrequency ablation in combination with transarterial chemoembolization for hepatocellular carcinoma | 10.3748/wjg.v19.i24.3872 | SR | WOS:000343926500012 |
| Radiofrequency (thermal) ablation versus no intervention or other interventions for hepatocellular carcinoma | 10.1002/14651858.CD003046.pub3 | SR | WOS:000329188300014 |
| EASL-EORTC Clinical Practice Guidelines: Management of hepatocellular carcinoma | | SR | WOS:000301849900001 |
| Retractions in the medical literature: how many patients are put at risk by flawed research? | 10.1136/jme.2011.043133 | SR | WOS:000296202900011 |
| ACE inhibitors, angiotensin receptor blockers and direct renin inhibitors in combination: a review of their role after the ONTARGET trial | 10.1185/03007990903152045 | SR | WOS:000269930900022 |
| ACEI/ARB therapy for IgA nephropathy: a meta analysis of randomised controlled trials | 10.1111/j.1742-1241.2009.02038.x | SR | WOS:000266025900013 |
| Role of ARBs in the Blood Hypertension Therapy and Prevention of Cardiovascular Events | 10.2174/138945009787122897 | SR | WOS:000263290800003 |
| Inhibiting the renin-angiotensin system: Why and in which patients | 10.1111/j.1745-7599.2008.00374.x | SR | WOS:000262227300009 |
| Change in proteinuria after adding aldosterone Blockers to ACE inhibitors or angiotensin receptor Blockers in CKD: A systematic review | 10.1053/j.ajkd.2007.10.040 | SR | WOS:000254799600006 |
| Guidelines 2007 Guidelines for the treatment of hypertension by the Committee for the drafting of the Guidelines of the European Society of Hypertension (ESH) and the European Society of Cardiology (ESC) | | CPG | WOS:000444204600001 |
| The 2007 Canadian Hypertension Education Program recommendations for the management of hypertension: Part 2 - therapy | 10.1016/S0828-282X(07)70798-5 | SR | WOS:000247448700004 |
| KDOQI clinical practice guidelines and clinical practice recommendations for diabetes and chronic kidney disease | 10.1053/j.ajkd.2006.12.004 | SR | WOS:000310508100041 |
| Role of angiotensin-receptor blockers in the prevention of cardiovascular risk: Clinical guidelines | 10.1007/978-88-470-0636-2_52 | CPG | WOS:000248938900052 |
| Japanese Society of Hypertension Guidelines for the Management of Hypertension (JSH 2004) | 10.1291/hypres.29.s1 | CPG | WOS:000241900100001 |
| Combination therapy with an angiotensin receptor blocker and an ACE inhibitor in proteinuric renal disease: A systematic review of the efficacy and safety data | 10.1053/j.ajkd.2006.04.077 | SR | WOS:000238826200002 |
| Angiotensin II antagonists - therapeutic benefits spanning the cardiovascular disease continuum from hypertension to heart failure and diabetic nephropathy | 10.1185/030079905X75041 | SR | WOS:000234751400001 |
| 2005 Spanish guidelines in diagnosis and treatment of arterial hypertension | 10.1157/13076402 | CPG | WOS:000230372100008 |
| Systematic review of combined angiotensin-converting enzyme inhibition and angiotensin receptor blockade in hypertension | 10.1161/01.HYP.0000161880.59963.da | SR | WOS:000228730300013 |
| Effects of angiotensin converting enzyme inhibitors and angiotensin II receptor antagonists on mortality and renal outcomes in diabetic nephropathy: systematic review | 10.1136/bmj.38237.585000.7C | SR | WOS:000224472800018 |
| Management of sexual dysfunction due to central nervous system disorders: a systematic review | 10.1111/bju.13055 | SR | WOS:000353228400011 |
| Treating Erectile Dysfunction and Central Neurological Diseases with Oral Phosphodiesterase Type 5 Inhibitors. Review of the Literature | 10.1111/j.1743-6109.2011.02615.x | SR | WOS:000302071600005 |
| The Movement Disorder Society Evidence-Based Medicine Review Update: Treatments for the Non-Motor Symptoms of Parkinson's Disease | 10.1002/mds.23884 | SR | WOS:000296611500003 |
| Surgical treatment of fibroids for subfertility | 10.1002/14651858.CD003857.pub4 | SR | WOS:000239141400024 |
| Hysteroscopy for treating subfertility associated with suspected major uterine cavity abnormalities | 10.1002/14651858.CD009461.pub4 | SR | WOS:000455302700038 |
| Efficacy of hysteroscopy in improving reproductive outcomes of infertile couples: a systematic review and meta-analysis | 10.1093/humupd/dmw008 | SR | WOS:000379742800006 |
| Hysteroscopy for Infertile Women: A Review | 10.1016/j.jmig.2014.12.163 | SR | WOS:000351481200008 |
| Surgical Interventions to Improve In Vitro Fertilization Outcomes: A Systematic Review of the Literature | 10.1089/gyn.2012.0092 | SR | WOS:000218064400001 |
| AAGL Practice Report: Practice Guidelines for the Diagnosis and Management of Submucous Leiomyomas | 10.1016/j.jmig.2011.09.005 | CPG | WOS:000301465000006 |
| The place of myomectomy in woman of reproductive age | 10.1016/j.jgyn.2011.09.023 | SR | WOS:000298075400022 |
| Evidence-based review of enhancing postoperative recovery after breast surgery | 10.1002/bjs.7331 | SR | WOS:000286538400005 |
| Drugs for preventing postoperative nausea and vomiting | 10.1002/14651858.CD004125.pub2 | SR | WOS:000239141400031 |
| Serum zinc level and children's asthma: A systematic and meta-analysis review article | 10.22088/cjim.12.3.236 | SR | WOS:000669710500001 |
| Pollution and respiratory disease: can diet or supplements help? A review | 10.1186/s12931-018-0785-0 | SR | WOS:000431270100001 |
| Paucity of evidence for a relationship between long-chain omega-3 fatty acid intake and chronic obstructive pulmonary disease: a systematic review | 10.1093/nutrit/nuv017 | SR | WOS:000363179000003 |
| Vitamins C and E for asthma and exercise-induced bronchoconstriction | 10.1002/14651858.CD010749.pub2 | SR | WOS:000338309000034 |
| Diet and Allergic Diseases among Population Aged 0 to 18 Years: Myth or Reality? | 10.3390/nu5093399 | SR | WOS:000328627500006 |
| Vitamin C for asthma and exercise-induced bronchoconstriction | 10.1002/14651858.CD010391.pub2 | SR | WOS:000326373200042 |
| K/DOQI clinical practice guidelines on hypertension and anti hypertensive agents in chronic kidney disease | 10.1053/j/ajkd.2004.03.004 | SR | WOS:000221669300001 |
| 2003 European society of hypertension - European Society of Cardiology guidelines for the management of arterial hypertension | 10.1097/01.hjh.0000059051.65882.32 | CPG | WOS:000183404100001 |
| Neurogenic Sexual Dysfunction Treatment: A Systematic Review | 10.1016/j.euf.2019.12.002 | SR | WOS:000558464000018 |
| Pharmacotherapy for Erectile Dysfunction: Recommendations From the Fourth International Consultation for Sexual Medicine (ICSM 2015) | 10.1016/j.jsxm.2016.01.016 | SR | WOS:000384727400001 |
| Intramedullary versus extramedullary internal fixation for unstable intertrochanteric fracture, a meta-analysis | 10.1016/j.aott.2018.02.009 | SR | WOS:000444010800011 |
| Nail or plate fixation for A3 trochanteric hip fractures: A systematic review of randomised controlled trials | 10.1016/j.injury.2018.05.017 | SR | WOS:000437364200013 |
| Comparing surgical interventions for intertrochanteric hip fracture by blood loss and operation time: a network meta-analysis | 10.1186/s13018-018-0852-8 | SR | WOS:000435965000001 |
| Internal fixation treatments for intertrochanteric fracture: a systematic review and meta-analysis of randomized evidence | 10.1038/srep18195 | SR | WOS:000366184100001 |
| Comparison of tip apex distance and cut-out complications between helical blades and lag screws in intertrochanteric fractures among the elderly: a meta-analysis | 10.1007/s00776-015-0770-0 | SR | WOS:000365421500016 |
| Circumcision devices versus standard surgical techniques in adolescent and adult male circumcisions | 10.1002/14651858.CD012250.pub2 | SR | WOS:000708167600001 |
| Applied techniques for putting pre-visit planning in clinical practice to empower patient-centered care in the pandemic era: a systematic review and framework suggestion | 10.1186/s12913-021-06456-7 | SR | WOS:000656260100002 |
| A review of question prompt lists used in the oncology setting with comparison to the Patient Concerns Inventory | 10.1111/ecc.12489 | SR | WOS:000423383300006 |
| Question Prompt Lists in health consultations: A review | 10.1016/j.pec.2015.05.015 | SR | WOS:000368332400002 |
| Improving subjective perception of personal cancer risk: systematic review and meta-analysis of educational interventions for people with cancer or at high risk of cancer | 10.1002/pon.3476 | SR | WOS:000337532400002 |
| A systematic review of interventions to facilitate ambulatory laparoscopic cholecystectomy | 10.1111/j.1477-2574.2011.00371.x | CPG | WOS:000295375500001 |
| Efficacy of physical activity interventions on psychological outcomes in refugee, asylum seeker and migrant populations: A systematic review and meta-analysis | 10.1016/j.psychsport.2021.101901 | SR | WOS:000709012600014 |
| Interventions for adults with a history of complex traumatic events: the INCiTE mixed-methods systematic review | 10.3310/hta24430 | SR | WOS:000569078200001 |
| PSYCHOLOGICAL INTERVENTION FOR POST-TRAUMATIC STRESS DISORDER COMORBID TO CHRONIC MUSCULOSKELETAL AND PRIMARY PAIN: A SYSTEMATIC REVIEW | | SR | WOS:000598576000003 |
| Efficacy and acceptability of psychosocial interventions in asylum seekers and refugees: systematic review and meta-analysis | 10.1017/S2045796019000027 | SR | WOS:000512676300006 |
| Chronic pain in refugees with posttraumatic stress disorder (PTSD): A systematic review on patients' characteristics and specific interventions | 10.1016/j.jpsychores.2018.07.014 | SR | WOS:000460190000015 |
| Schemas and coping strategies in cognitive-behavioral therapy for PTSD: A systematic review | 10.1016/j.ejtd.2018.09.005 | SR | WOS:000646066500005 |
| Psychosocial interventions for post-traumatic stress disorder in refugees and asylum seekers resettled in high-income countries: Systematic review and meta-analysis | 10.1371/journal.pone.0171030 | SR | WOS:000396161200075 |
| Interventions for treating persistent pain in survivors of torture | 10.1002/14651858.CD012051.pub2 | SR | WOS:000408828100023 |
| Chronic Pain and Cognitive Behavioral Therapy: An Integrative Review | 10.1177/0193945915615869 | SR | WOS:000374347600005 |
| Are multidisciplinary interventions multicultural? A topical review of the pain literature as it relates to culturally diverse patient groups | 10.1097/j.pain.0000000000000412 | SR | WOS:000378257600007 |
| Psychological, social and welfare interventions for psychological health and well-being of torture survivors | 10.1002/14651858.CD009317.pub2 | SR | WOS:000347646200035 |
| Primary Care Management of Non-English-Speaking Refugees Who Have Experienced Trauma A Clinical Review | 10.1001/jama.2013.8788 | SR | WOS:000322786400024 |
| Psychological therapies for the management of chronic pain (excluding headache) in adults | 10.1002/14651858.CD007407.pub3 | SR | WOS:000312199800004 |
| Comparison of early and delayed removal of dressing following primary closure of clean and contaminated surgical wounds: A systematic review and meta-analysis of randomized controlled trials | 10.3892/etm.2020.8591 | SR | WOS:000528950100006 |
| Patient education for preventing recurrence of venous leg ulcers: a systematic review | | SR | WOS:000513010100002 |
| Role of Honey in Topical and Systemic Bacterial Infections | 10.1089/acm.2017.0017 | SR | WOS:000422937900004 |
| A Systematic Review and Meta-Analysis of Nutritional Supplementation in Chronic Lower Extremity Wounds | 10.1177/1534734616674624 | SR | WOS:000392883300003 |
| Honey: A Therapeutic Agent for Disorders of the Skin | 10.5195/cajgh.2016.241 | SR | WOS:000407876300005 |
| A systematic review and meta-analysis of dressings used for wound healing: the efficiency of honey compared to silver on burns | 10.1080/10376178.2016.1171727 | SR | WOS:000375908400002 |
| Debridement for venous leg ulcers | 10.1002/14651858.CD008599.pub2 | SR | WOS:000209933500019 |
| Honey as a topical treatment for wounds | 10.1002/14651858.CD005083.pub4 | SR | WOS:000375542100010 |
| Comparative effectiveness of advanced wound dressings for patients with chronic venous leg ulcers: A systematic review | 10.1111/wrr.12151 | SR | WOS:000332835400187 |
| Honey in modern wound care: A systematic review | 10.1016/j.burns.2013.06.014 | SR | WOS:000329265700002 |
| Evidence for Clinical Use of Honey in Wound Healing as an Anti-bacterial, Anti-inflammatory Anti-oxidant and Anti-viral Agent: A Review | 10.17795/jjnpp-9487 | SR | WOS:000421138200002 |
| Honey and Wound Healing An Overview | 10.2165/11538930-000000000-00000 | SR | WOS:000291277800005 |
| Ketamine in adult cardiac surgery and the cardiac surgery Intensive Care Unit: An evidence-based clinical review | 10.4103/0971-9784.154478 | SR | WOS:000218777000015 |
| NMDA Receptor Antagonists, Gabapentinoids, alpha-2 Agonists, and Dexamethasone and Other Non-Opioid Adjuvants: Do They Have a Role in Plastic Surgery? | 10.1097/PRS.0000000000000703 | SR | WOS:000347249600011 |
| Does Intraoperative Ketamine Attenuate Inflammatory Reactivity Following Surgery? A Systematic Review and Meta-Analysis | 10.1213/ANE.0b013e3182662e30 | SR | WOS:000309490500032 |

SR, systematic review; CPG, clinical practice guideline
